# Genome-wide analysis of heart failure yields insights into disease heterogeneity and enables prognostic prediction in the Japanese population

**DOI:** 10.1101/2024.11.14.24317249

**Authors:** Nobuyuki Enzan, Kazuo Miyazawa, Satoshi Koyama, Ryo Kurosawa, Hirotaka Ieki, Hiroki Yoshida, Fumie Takechi, Masashi Fukuyama, Ryosuke Osako, Kohei Tomizuka, Xiaoxi Liu, Kouichi Ozaki, Yoshihiro Onouchi, BioBank Japan Project, Koichi Matsuda, Yukihide Momozawa, Hiroyuki Aburatani, Yoichiro Kamatani, Takanori Yamaguchi, Akazawa Hiroshi, Koichi Node, Patrick T. Ellinor, Michael G. Levin, Scott M. Damrauer, Benjamin F. Voight, Jacob Joseph, Yan V. Sun, Chikashi Terao, Toshiharu Ninomiya, Issei Komuro, Kaoru Ito

**Affiliations:** Laboratory for Cardiovascular Genomics and Informatics, RIKEN Center for Integrative Medical Sciences, Yokohama, Japan; Cardiovascular Research Center, Massachusetts General Hospital, Boston, MA, USA; Cardiovascular Disease Initiative, The Broad Institute of MIT and Harvard, Cambridge, MA, USA; Program in Medical and Population Genetics, Broad Institute of Harvard and MIT, Cambridge, MA, USA; Department of Genetics, Stanford University School of Medicine, Stanford, USA; Department of Cardiovascular Medicine, Graduate School of Medicine, The University of Tokyo, Tokyo, Japan; Department of pediatrics Graduate School of Medical and Pharmaceutical Sciences, Chiba University, Chiba, Japan; Department of Cardiovascular Medicine, Saga University, Saga, Japan; Laboratory for Statistical and Translational Genetics, RIKEN Center for Integrative Medical Sciences, Kanagawa, Japan; Medical Genome Center, Research Institute, National Center for Geriatrics and Gerontology, Obu, Japan; Department of Public Health, Chiba University Graduate School of Medicine, Chiba, Japan; Laboratory of Clinical Genome Sequencing, Department of Computational Biology and Medical Sciences, Graduate School of Frontier Sciences, The University of Tokyo, Tokyo, Japan; Laboratory for Genotyping Development, RIKEN Center for Integrative Medical Sciences, Kanagawa, Japan; Genome Science Division, Research Center for Advanced Science and Technology, The University of Tokyo, Tokyo, Japan; Laboratory of Complex Trait Genomics, Department of Computational Biology and Medical Sciences, Graduate School of Frontier Sciences, The University of Tokyo, Tokyo, Japan; Demoulas Center for Cardiac Arrhythmias, Massachusetts General Hospital, Boston, MA, USA; Division of Cardiovascular Medicine, Perelman School of Medicine, University of Pennsylvania, Philadelphia, PA, USA; Corporal Michael J. Crescenz VA Medical Center, Philadelphia, PA, USA; Department of Surgery, University of Pennsylvania Perelman School of Medicine, Philadelphia, PA, USA; Department of Genetics, University of Pennsylvania Perelman School of Medicine, Philadelphia, PA, USA; Department of Systems Pharmacology and Translational Therapeutics, University of Pennsylvania Perelman School of Medicine, Philadelphia, PA, USA; Institute of Translational Medicine and Therapeutics, University of Pennsylvania Perelman School of Medicine, Philadelphia, PA, USA; Veterans Affairs Providence Healthcare System, Providence, RI, USA; Brown University, Providence, RI, USA; VA Atlanta Health Care System, Decatur, GA, USA; Emory University Rollins School of Public Health, Atlanta, GA, USA; Department of Epidemiology and Public Health, Graduate School of Medical Sciences, Kyushu University.; Department of Frontier Cardiovascular Science, Graduate School of Medicine, The University of Tokyo, Tokyo, Japan; International University of Health and Welfare.

## Abstract

To understand the genetic basis of heart failure (HF) in the Japanese population, we performed genome-wide association studies (GWASs) comprising 16,251 all-cause HF cases, 4,254 HF with reduced ejection fraction cases, 7,154 HF with preserved ejection fraction cases, and 11,122 non-ischemic HF cases among 213,828 individuals and identified five novel loci. A subsequent cross-ancestry meta-analysis and multi-trait analysis of the GWAS data identified 19 novel loci in total. Among these susceptibility loci, a common non-coding variant in *TTN* (rs1484116) was associated with reduced cardiac function and worse long-term mortality. We leveraged the HF meta-GWASs along with cardiac function-related GWASs to develop a polygenic risk score (PRS) for HF. The PRS successfully identified early-onset HF and those with an increased risk of long-term HF mortality. Our results shed light on the shared and distinct genetic basis of HF between Japanese and European populations and improve the clinical value of HF genetics.

## Introduction

Heart failure (HF) is increasing in prevalence and incidence^1^. A previous genome-wide association study (GWAS) of all-cause HF identified 47 risk loci^2^. Considering that GWAS of coronary artery disease (CAD) yielded 175 susceptibility loci^3^ and GWAS of atrial fibrillation (AF) yielded 150 loci^4^, the number of loci for HF was lower than expected even when using multi-trait analysis of GWAS (MTAG) that jointly analyzes GWAS summary statistics of multiple related traits, boosting statistical power by leveraging information across traits. This is partly because HF is a heterogeneous syndrome resulting from multiple etiologies. GWAS of HF with reduced ejection fraction (HFrEF) and HF with preserved ejection fraction (HFpEF) was performed to address this heterogeneity^5^. This study identified additional susceptibility loci that had not been found in the all-cause HF GWAS. Given this fact, analysis of HF subtypes should be useful to better understand the genetic architecture for HF. On the other hand, since the vast majority of these HF-GWASs have been performed in European populations, the genetic pathophysiology of HF in non-European populations is not well understood. Additionally, it is difficult to apply polygenic risk scores (PRSs) derived from such GWASs to non-European populations.

Therefore, in the present study, we performed a large-scale Japanese GWAS to explore the genetic architecture of all-cause HF and HF subtypes: HFrEF, HFpEF, and non-ischemic HF (NIHF), followed by a cross-ancestry meta-analysis. Further, we investigated potential causal genes at the identified HF-associated loci by integrating several prioritization methods to characterize the underlying mechanism, clarify the link with cardiomyopathy, and propose gene-drug interactions relevant to HF. We also evaluated the impact of common variants in established cardiomyopathy genes on HF phenotypes and long-term mortality. Subsequently, we identified a *TTN* common variant, which affected HF severity and mortality rate. Additionally, we developed a PRS derived from the cross-ancestry meta-analysis of each HF subtype along with HF-related phenotypes and demonstrated the ability of the HF-PRS to predict the early onset of HF and stratify the long-term mortality, which may provide evidence for the clinical utility of HF-PRS and lay the foundation for the realization of precision medicine in HF.

## Results

### GWAS revealed a Japanese-specific genetic architecture of HF

An overview of the study design is shown in **Fig. 1**. We performed Japanese case-control GWASs to investigate the association of up to 7,974,473 common genetic variants in the autosomes (minor allele frequency (MAF) > 1%) with the risk of four HF phenotypes. 16,251 HF cases were identified, including 4,254 cases of HFrEF, 7,154 cases of HFpEF, and 11,122 cases of NIHF (**Fig. 1**). Cases of all-cause HF, HFrEF, and HFpEF were compared with 197,577 controls. For the NIHF analysis, CAD cases were excluded from control samples, and 171,995 controls were used. Sample overlaps between each HF phenotype are shown in **Supplementary Fig. 1a**. The mean LV ejection fraction (LVEF) was 48.94% for all-cause HF, 28.74% for HFrEF, 62.08% for HFpEF, and 50.86% for NIHF (**Supplementary Table 1**). These GWASs identified 18 genome-wide significant loci in total, of which five were previously unreported **(Fig. 2a**, **Supplementary Fig. 1b, Supplementary Fig. 1c, Supplementary Table 2**, and **Supplementary Datasets**). The estimated heritability for each HF phenotype is described in **Supplementary Notes 1**. We assessed the 62 previously reported HF loci and confirmed that their effects were mostly concordant with our Japanese GWAS (**Supplementary Notes 2**, **Supplementary Fig. 2, Supplementary Table 3**). Additionally, while the reproducibility of the novel loci was obtained in both internal and external replications, two of the novel loci were specific to the Japanese population and were not found in Europeans (**Supplementary Notes 3**, **Supplementary Fig. 3a, b, c, d and Supplementary Table 4, 5**).

**Fig. 1.**
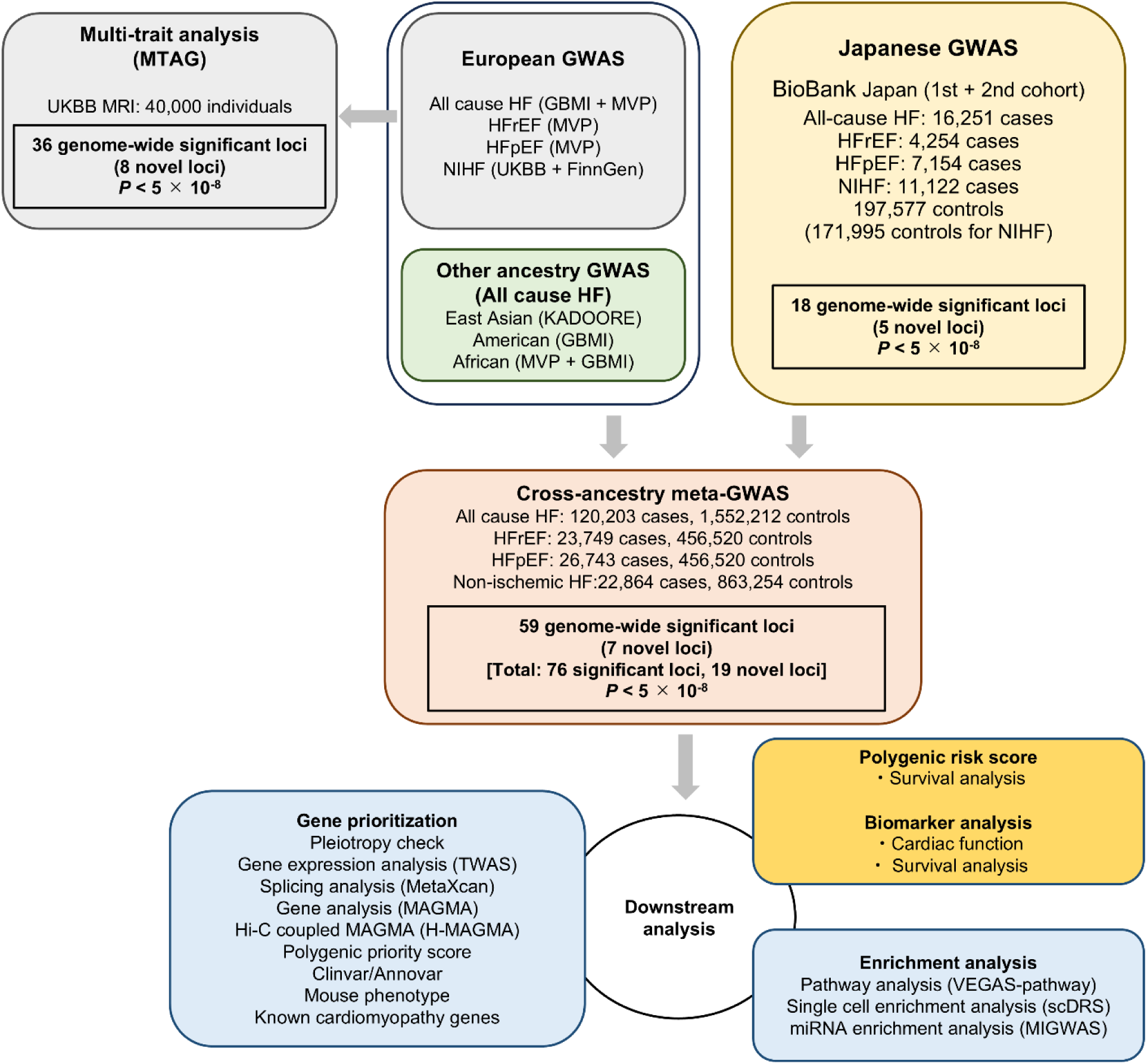
Overview of the study design. Flowchart of the study, which encompasses the Japanese GWAS with the BioBank Japan, a trans-ancestry meta-analysis with large-scale European and other population GWAS followed by the downstream analysis. GWAS, genome wide association study; HF, heart failure; HFpEF, heart failure with preserved ejection fraction; HFrEF, heart failure with reduced ejection fraction; NIHF; non-ischemic heart failure; TWAS, transcriptome-wide association study.

**Fig. 2.**
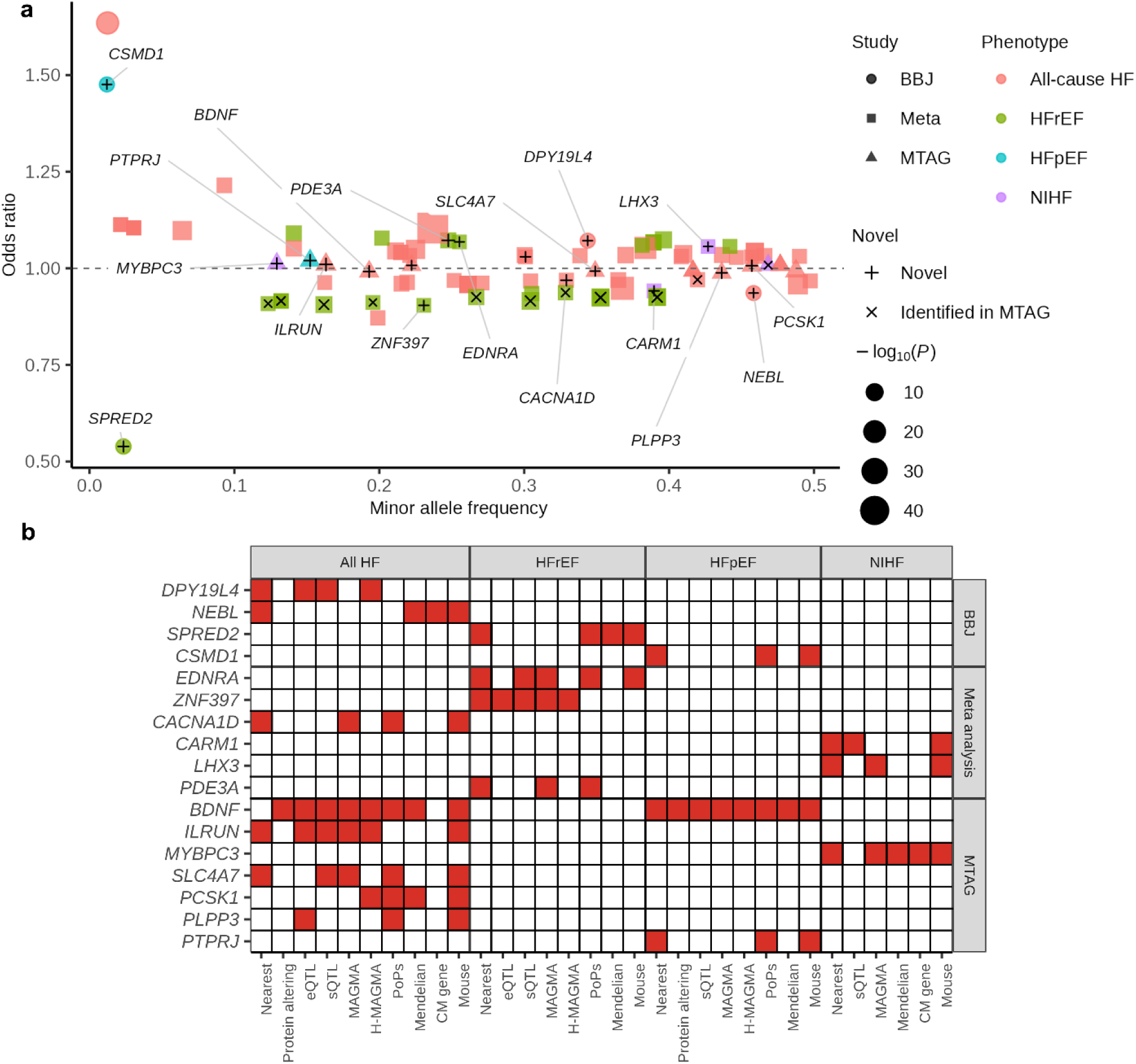
Prioritized genes. **(a)** The odds ratios for HF development of the independent signals in the Japanese GWAS, cross-ancestry meta-analysis, and MTAG. The color of each point indicates the corresponding HF phenotype. The size of each point indicates -log_10_(P value). The shape of each point indicates the methods of analysis. The marker inside each point indicates novelty status. **(b)** Prioritized genes for each HF phenotype according to the types of analyses. *CACNA1D* was prioritized in both BBJ and cross-ancestry meta-analysis and is shown in the “Meta-analysis” row. BBJ, BioBank Japan; HF, heart failure; HFpEF, heart failure with preserved ejection fraction; HFrEF, heart failure with reduced ejection fraction; Meta, meta-analysis; MTAG, multi-trait analysis of genome-wide association study; NIHF, non-ischemic heart failure.

To identify HF-associated variants independent of the lead variant at each locus, we performed a stepwise conditional analysis, in which three independent variants (locus-wide *P* < 5.0 × 10^-6^) were additionally detected, increasing the total number of HF-associated signals to 21 (**Supplementary Table 6**). Among these, the frequencies of rs893363 (Alternative allele frequency; EAS 96.2% vs. EUR 61.9%, β = −0.167 in NIHF) and rs4307025 (Alternative allele frequency; EAS 73.0% vs. EUR 28.0%, β = 0.0912 in NIHF) differed significantly between East Asians and Europeans.

To assess the pleiotropic effects, we used the lead variant in each locus as a proxy and assessed its effect sizes in Japanese GWAS of cardiovascular-related phenotypes^6^. The novel variant rs35593046 found in all-cause HF (β = −0.0630) /NIHF (β = −0.0767) had a protective role against high blood pressure (**Supplementary Fig. 4a** and **Supplementary Table 7**). It is the intronic variant of *CACNA1D* and is much more frequently observed in the East Asian population than in European populations (53.8% vs. 26.8%).

### Common variants in cardiomyopathy genes were responsible for HF development

We then sought to prioritize potential causal genes at the identified HF-associated loci. First, of the 310 variants in LD (r^2^ > 0.8) with 18 lead variants, 6 missense variants were observed (**Supplementary Table 8**). Among loci previously reported only in the multi-trait analysis of GWAS (MTAG), we found missense variants rs11718898, rs2305398, and rs3732678 in the *CAND2* gene, three of which were in high LD. *CAND2* has been reported to link mTOR signaling to pathological cell growth leading to cardiac remodeling^7^. A missense variant rs2627043 found in the HFrEF GWAS encodes *TTN*, one of the well-known cardiomyopathy genes. Notably, the allele frequency was much higher in the East Asian population than in European populations (64.7% vs. 20.1%). We also found the East Asian-specific missense variant rs671 encoding *ALDH2,* which has been reported to be an HF-related variant in the BBJ 1^st^ cohort^6^.

Next, we assessed whether the lead variants we identified were functioning as an expression quantitative trait locus (eQTL) or a splicing quantitative trait locus (sQTL) using GTEx version 8 data. Among the lead variants in novel loci, rs6471480 was found to be an eQTL in subcutaneous adipose tissue (**Supplementary Fig. 4c** and **Supplementary Table 9**) and an sQTL in the left atrial appendage (**Supplementary Fig. 4d** and **Supplementary Table 10**) for the *DPY19L4* gene. Additionally, all of the five HFrEF-related variants, previously identified only in MTAG with cardiac measurements^2^, were revealed as eQTL in the left ventricle. This result may suggest that MTAG in the previous study conferred additional statistical power to discover these variants by integrating cardiac function-related traits.

We conducted gene-based analyses, MAGMA^8^ and H-MAGMA^9^, utilizing promoter capture Hi-C in cardiovascular-related tissues^10^. Among novel loci, *DPY19L4* reached statistical significance (FDR < 0.05) in the aorta and adrenal gland using H-MAGMA but not MAGMA (**Supplementary Fig. 4e** and **Supplementary Table 11-12**). Given that the aorta and adrenal gland are known to be involved in the pathophysiology of hypertension, this is consistent with the association of *DPY19L4* with the use of a calcium channel blocker, one of the antihypertensive drugs^6^.

Furthermore, related to the HF loci identified in our Japanese GWAS, we assessed nearby genes containing variants classified in ClinVar as having pathogenic evidence for HF-relevant monogenic disorders, knock-out mouse phenotype, and known cardiomyopathy genes^11^. *NEBL*, located within 500kb of the lead variant rs7075837, was reported to harbor pathogenic variants responsible for primary dilated cardiomyopathy according to the ClinVar database (**Supplementary Table 13**). *NEBL* was also one of the known cardiomyopathy genes (**Supplementary Table 14**), and *Nebl* knock-out mice showed dilatation of the left atrium and ventricle (**Supplementary Table 15**). *SPRED2* harbors pathogenic variants responsible for Noonan syndrome, which causes hypertrophic cardiomyopathy (**Supplementary Table 13**). Knock-out of *Spred2* also causes dilatation of the heart (**Supplementary Table 15)**. Knock-out of *Csmd1* showed obesity and abnormal glucose metabolism (**Supplementary Table 15)**. Polygenic Priority Score (PoPS) was recently introduced to estimate responsible genes using various gene features, such as cell-type-specific gene expression and biological pathways. We thus used PoPS to choose the top two likely genes in each locus. As a result, *SPRED2* and *CSMD1* were also prioritized (**Supplementary Table 16**).

Taken together, *DPY19L4*, *CSMD1*, *SPRED2*, and *NEBL* were prioritized by at least three out of 10 predictors as Japanese-specific HF-related genes (**Fig. 2a, 2b,** and **Supplementary Table 17**). Of note, common variants in cardiomyopathy genes *NEBL* were also responsible for HF development. *NEBL* encodes the cardiac Z-disk protein nebulette, and rare variants in this region cause dilated, hypertrophic, and LV non-compaction cardiomyopathy^12^.

*DPY19L4* was likely to act on arteries to cause hypertension based on pleiotropic effects and H-MAGMA results. This interpretation was supported by the fact that quercetin, which attenuated atherosclerosis via modulating oxidized LDL-induced endothelial cellular senescence, downregulated *DPY19L4* in human aortic endothelial cells^13^. Based on knockout phenotypes, *CSMD1* is likely to cause HF through metabolic disorders, consistent with a previous study^14^.

*SPRED2* loss-of-function induced defects in convergence and extension cell movements leading to cardiac defects and Noonan-like syndrome^15^. Aside from novel loci, *AOPEP,* previously reported in Japanese all-cause HF GWAS^6^, was also associated with HFrEF, HFpEF, and NIHF and the Japanese-specific signal (**Supplementary Fig. 5**). Given that *AOPEP* cleaves angiotensin III to angiotensin IV, a bioactive peptide of the renin-angiotensin pathway, it is likely that *AOPEP* is responsible for HF development through hypertension.

### Cross-ancestry meta-analysis identified seven novel loci for HF

To improve the statistical power to detect further genetic associations with HF, we conducted cross-ancestry meta-analyses by combining the current Japanese GWAS (BBJ), along with European^2^, Chinese^16^, American^17^, and African^2^ all-cause HF GWAS, European HFrEF and HFpEF GWAS^5^, and the UK biobank^18^ and FinnGen (data release 9) NIHF GWAS (**Supplementary Table 18** and **Supplementary Notes 4**). These analyses identified 58 HF-associated loci with genome-wide significance (*P* < 5.0 × 10^-8^; **Supplementary Fig. 6, Supplementary Fig. 7a, Supplementary Table 19**, and **Supplementary Datasets**). Of these loci, seven have not been reported previously, including one novel locus detected in the current Japanese GWAS (*CACNA1D* locus). In total, we identified 11 novel loci through the current Japanese GWAS and cross-ancestry meta-analysis.

In internal replication, all the newly identified loci were nominally significant both in East Asian and European populations (**Supplementary Fig. 7b** and **Supplementary Table 20**). rs35593046 (*CACNA1D*), rs7659823 (*EDNRA*), rs5823966 (*ZNF397*), and rs1541596 (*CARM1*) were much more frequently observed in the East Asian population (**Supplementary Fig. 7c** and **Supplementary Table 20**). Replication analyses of three lead variants in HFrEF and HFpEF using independent datasets showed concordant results with the same effect direction (**Supplementary Notes 5**, **Supplementary Figure 7d, e**, and **Supplementary Table 21**).

### Genes related to arrhythmia, metabolism, and heart morphology were prioritized in the cross-ancestry meta-analyses

Of the 1,621 variants in LD (*r*^2^ > 0.8) with 75 lead variants in any of the HF subtypes, 10 missense variants and one stop-gain variant were observed (**Supplementary Table 22**). We found an additional missense variant rs2305397 in the *CAND2* gene.

We performed a transcriptome-wide association study (TWAS) and splicing-TWAS using the identified loci in the cross-ancestry meta-analyses and the GTEx data. Among novel loci, *ZNF397* was prioritized in the tibial artery by TWAS (**Supplementary Fig. 8a** and **Supplementary Table 23**). As for loci previously found only in MTAG, *MTSS1*, *PROM1*, and *SPATA24* were prioritized in the left ventricle or left atrial appendage and *CCDC136* was prioritized in the tibial artery. The splicing-TWAS showed that *ZNF397* (left ventricle) and *EDNRA* (left ventricle and left atrial appendage) were prioritized among the novel loci (**Supplementary Fig. 8b** and **Supplementary Table 24**). *TTN*, *SMARCB1*, *SPATA24*, *EFCAB13*, *CCDC136*, *CAND2*, and *CARM1* were prioritized in the left ventricle or left atrial appendage. Gene-based analysis by MAGMA showed that *CACNA1D*, *LHX3*, *EDNRA*, and *ZNF397* reached statistical significance (**Supplementary Fig. 8c** and **Supplementary Table S25** and **S26**).

Among the novel loci, no gene harbored pathogenic variants responsible for cardiovascular-related phenotypes according to the ClinVar database (**Supplementary Table 27**). Based on genetic knockout phenotypes in mice, *Cacna1d* deletion caused arrhythmia (**Supplementary Table 28**) and was prioritized by PoPS (**Supplementary Table 29**). Knockout of *Ednra* showed abnormal heart and aortic morphology (**Supplementary Table 28)** partly because *Ednra* was associated with heart development^19^. Deletion of *Carm1* caused dilatation of the left atrium, and deletion of *Lhx3* caused decreased body weight (**Supplementary Table 28)**.

In the cross-ancestry meta-analyses, *EDNRA*, *ZNF397*, *CACNA1D*, *CARM1*, *LHX3*, and *PDE3A* were prioritized by at least three predictors (**Fig. 2a, 2b,** and **Supplementary Table 30**). Among these six genes, *EDNRA* and *PDE3A* were linked to existing medications (refer to **Candidate drugs linked to disease susceptibility loci** section in the main text). No prioritized gene was found associated with rs10851802 (**Supplementary Notes 6)**.

### The HF-related common variant in the cardiomyopathy gene *MYBPC3* was identified in MTAG

To further enhance the statistical power of HF GWAS, we conducted MTAG^20^. First, we assessed the genetic correlation between HF and cardiac MRI parameters for the left ventricle^11^, left atrium^21^, right ventricle, right atrium^22^, left ventricular mass^23^, and fibrosis^24^. Because left ventricular mass was strongly correlated with HF in all of the HF subtypes (**Supplementary Fig. 9a** and **Supplementary Table 31**), we integrated HF GWAS and left ventricular mass GWAS by MTAG. We then found an additional eight novel loci (**Supplementary Fig. 9b, 9c, Supplementary Table 32**, and **Supplementary Datasets**). Of the 1,597 variants in LD (*r*^2^ > 0.8) with 36 lead variants, seven missense variants were observed (**Supplementary Table 33**). Among the novel loci, rs6265 is a missense variant for *BDNF* (**Supplementary Table 33**). TWAS showed that *BDNF* and *ILRUN* were prioritized in the left ventricle and atrial appendage, and *PLPP3* in the fibroblasts (**Supplementary Fig. 10a** and **Supplementary Table 34**). Splicing-TWAS showed that *ILRUN* (left ventricle and atrial appendage), *BDNF* (coronary artery/aorta/fibroblasts), and *SLC4A7* (fibroblasts) were prioritized (**Supplementary Figure 10b** and **Supplementary Table 35**). *SLC4A7*, *MYBPC3*, *BDNF*, and *PCSK1* reached statistical significance in gene-based analyses (**Supplementary Fig. 10c** and **Supplementary Table 36-37**).

Combined with known Mendelian genes (**Supplementary Table 38**), mouse phenotypes (**Supplementary Table 39**), and PoPS (**Supplementary Table 40**), *BDNF*, *MYBPC3*, *SLC4A7*, *ILRUN*, *PCSK1*, *PTPRJ*, and *PLPP3* were prioritized (**Fig. 2a**, **2b**, **Supplementary Table 41**). Notably, common variants in one of the known cardiomyopathy genes *MYBPC3* were responsible for HF development as well as *TTN*, *NEBL* and *BAG3*. Other prioritized genes are described in **Supplementary Notes 7**.

Here, comparing the disease susceptibility loci detected by the Japanese GWAS, European GWAS, cross-ancestry meta-analyses, and MTAG, we found that there were common and different genetic effects between the East Asian and European populations. Among novel loci found in BBJ-specific or cross-ancestry meta-analyses, *AOPEP*, *CSMD1*, *DPY19L4*, and *SPRED2* were significant only in the East Asian population (**Supplementary Fig. 4** and **Supplementary Fig. 11**). On the other hand, *HLA-B*, *APOH*, *CFL2*, *HSD17B12*, *UBA7* were European specific loci (**Supplementary Fig. 11**).

### Distinct mechanism of each HF phenotype in cross-ancestry analysis

To compare the mechanisms of each HF phenotype, we conducted a pathway analysis and found that cellular senescence, cyclin-dependent kinase-related pathway, miRNAs involved in DNA damage responses, signaling events mediated by prolactin, Tie2 mediated signaling, and Notch signaling pathway were enriched in all-cause HF (**Fig. 3** and **Supplementary Table 42**). Notch signaling was also enriched in NIHF. From a clinical perspective, miRNAs involved in DNA damage response may be attractive targets to improve cancer patient outcomes in chemotherapy-related cardiomyopathy, which may be induced by DNA-damaging chemotherapy. The prolactin inhibitor bromocriptine had higher odds of left ventricular function recovery in peripartum cardiomyopathy, one of the leading causes of maternal mortality^25^. Tie2-mediated signaling is not only required for normal vascular development but also is required for ventricular chamber formation ^26^. Notch is associated with heart development and regulates cardiac regenerative processes^27^. Aside from cyclin-dependent kinase-related pathways, sarcomere-related pathways including Z-disc, I-band, and actin-binding were enriched in HFrEF (**Fig. 3** and **Supplementary Table 42**). Cardiac cell development/differentiation was enriched in NIHF. On the other hand, no specific pathway was enriched in HFpEF.

**Fig. 3.**
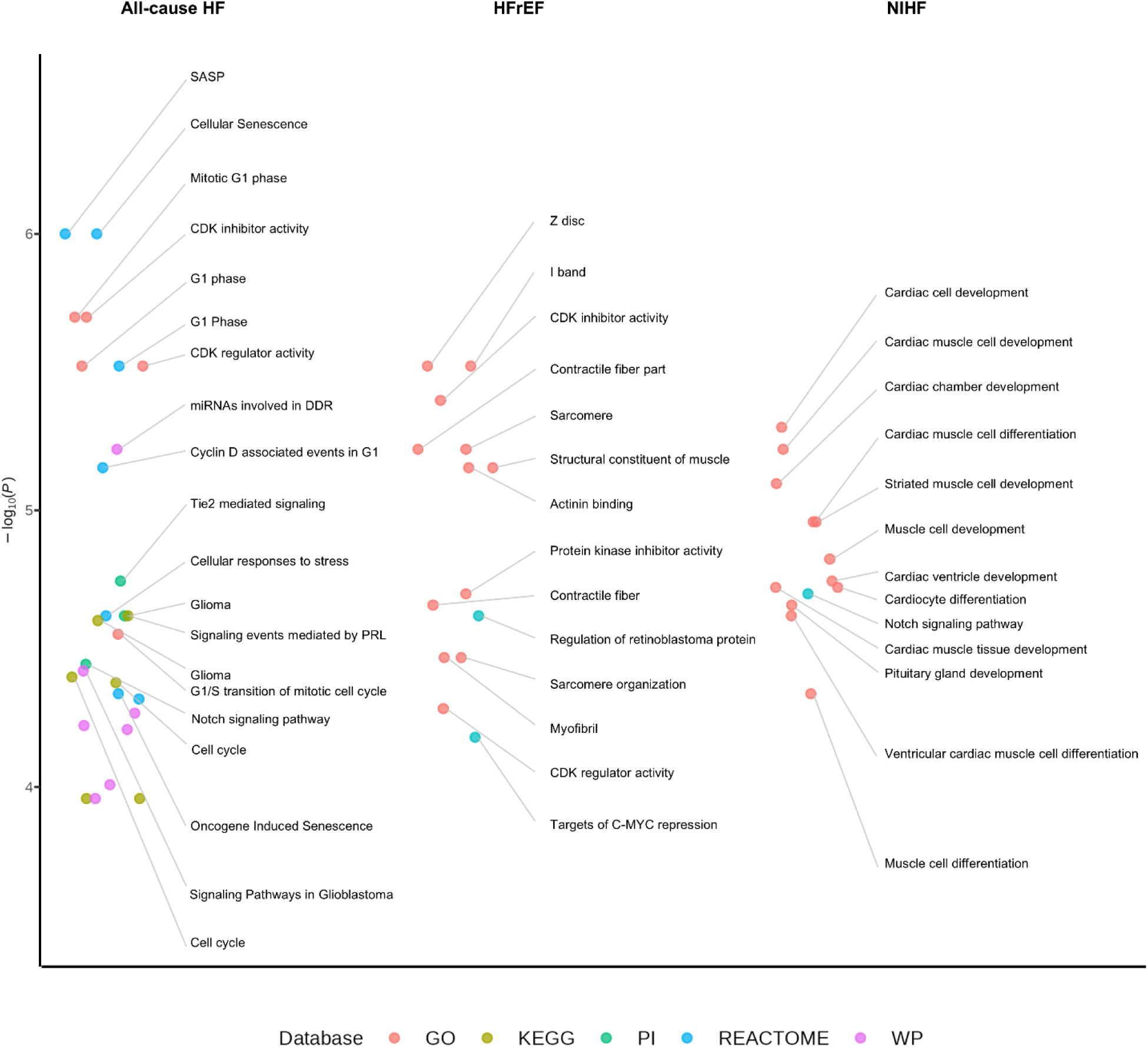
Pathway analysis for each HF phenotype. Enriched terms (FDR < 0.05) for each HF phenotype from GO, KEGG, PI, Reactome, and WP. Up to the top 20 enriched terms in each HF phenotype gene set are labeled. The color of the dots indicates the pathway. There was no enriched pathway for HFpEF. GO, Gene Ontology; KEGG, Kyoto Encyclopedia of Genes and Genomes; PI, Pathway Interaction database; WP, Wiki Pathways.

Additionally, cell type-specific enrichment analysis with gene expression and epigenetic marks showed that H3K4me1 in the fetal heart was enriched in HFrEF, but the other three HF phenotypes did not show any particular cell type enrichment (**Supplementary Table 43**). Taken together, despite the smaller sample size, HFrEF showed strong enrichment in the heart, especially in the sarcomere.

With miRNA enrichment analysis using MIGWAS, HFrEF showed significant enrichment for the miRNA-target gene network (**Supplementary Figure 12a**). Out of eight candidate miRNA-gene pairs (**Supplementary Table 44**), HSPB7 was also significant in TWAS analysis (**Supplementary Table 34**), suggesting that hsa-mir-4728-*HSPB7* was the most likely pair. *HSPB7* has been reported to be indispensable for heart development and associated with dilated cardiomyopathy^47^.

We conducted the cell type-specific analysis at single-cell resolution using scDRS^48^ with healthy, dilated cardiomyopathy, and hypertrophic cardiomyopathy datasets^49^. All four HF phenotypes showed significant enrichment for cardiomyocytes (**Supplementary Fig 12b** and **Supplementary Table 45**). Additionally, HFpEF showed enrichment for adipocyte and vascular smooth muscle cells^50, 51^, suggesting that HFpEF may have additional mechanisms beyond the other HF phenotypes.

### Candidate drugs linked to disease susceptibility loci

We curated drugs from DrugBank^28^ and Therapeutic Target Database^29, 30^ (**Supplementary Fig. 13**) for the prioritized genes from the Japanese GWAS, cross-ancestry meta-analyses, and MTAG. Our analysis supports epidemiological evidence that several medications cause or exacerbate HF. Given that knockout of *CACNA1D*, *EDNRA*, and *CDK6* in mice caused abnormal myocardial fiber physiology, abnormal heart ventricle morphology, and decreased myocardial fiber number, respectively, those inhibitors could worsen HF. Indeed, the previous meta-analysis suggested that dihydropyridine calcium channel blockers (nifedipine) can be a risk for HF^31^. The EDNRA inhibitor bosentan caused fluid retention in HF patients^32^. One of the CDK4/6 inhibitors, palbociclib, was associated with worse outcomes in patients who developed atrial fibrillation or HF^33^. All these outcomes were concordant with our genetic analyses.

Tirzepatide, one of the emerging antidiabetics^34, 35^, could be a potential drug repositioning candidate through the *GIPR* locus, which had a protective role against HF. As of now, tirzepatide has not been fully evaluated for its effects on cardiovascular outcomes, but at least it did not increase the risk of cardiovascular events^36^. Currently, a study of tirzepatide in participants with HFpEF (SUMMIT trial) is ongoing (NCT04847557).

### A *TTN* common variant plays an important role in HF outcomes

From the Japanese GWAS and cross-ancestry meta-analyses, we found lead variants associated with cardiomyopathy genes, *TTN*, *NEBL*, and *BAG3* (**Supplementary Table 14**). Given the nature of cardiomyopathy genes, we hypothesized that these variants by themselves could affect cardiac function and prognosis. Among those, the risk allele of *TTN* lead variant (rs1484116) was less frequent in the East Asian populations than in European populations (Risk allele frequency: EUR 0.800, EAS 0.315; **Fig. 4a**) according to the gnomAD database, while we observed the effect size of this variant was higher in Japanese than Europeans (**Fig. 4b**). First, we investigated the association between these variants and cardiac function, and we found that the lead variants of *TTN* (rs1484116) and *BAG3* (rs61870083) were significantly associated with lower left ventricular ejection fraction (*TTN*, effect size −1.16, 95% confidence interval (95%CI) −1.55 - −0.77, *P* = 7.80 × 10^-9^; *BAG3*, effect size −2.79, 95%CI −3.58 - −2.00, *P* = 3.88 × 10^-12^; **Fig. 4c**). We then repeated the same analysis in NIHF to exclude possible effects of CAD, and it yielded similar results (**Fig. 4c**). These variants of *TTN* and *BAG3* were also associated with lower LVEF in individuals without known cardiac diseases in UKBB cohorts (**Fig. 4c** right panel). Furthermore, we assessed the effects of these common variants on long-term HF mortality among non-HF subjects and HF subjects. The Kaplan-Meier estimates and Cox regression analysis demonstrated that the *TTN* lead variant (rs1484116) was significantly associated with worse outcomes in HF patients (HR 1.24, 95%CI 1.06-1.46, *P* = 8.12 × 10^-3^; **Fig. 4d** and **e**). These results suggest the importance of the *TTN* common variant in risk stratification of HF patients and this variant could be a novel potential biomarker for HF.

**Fig. 4.**
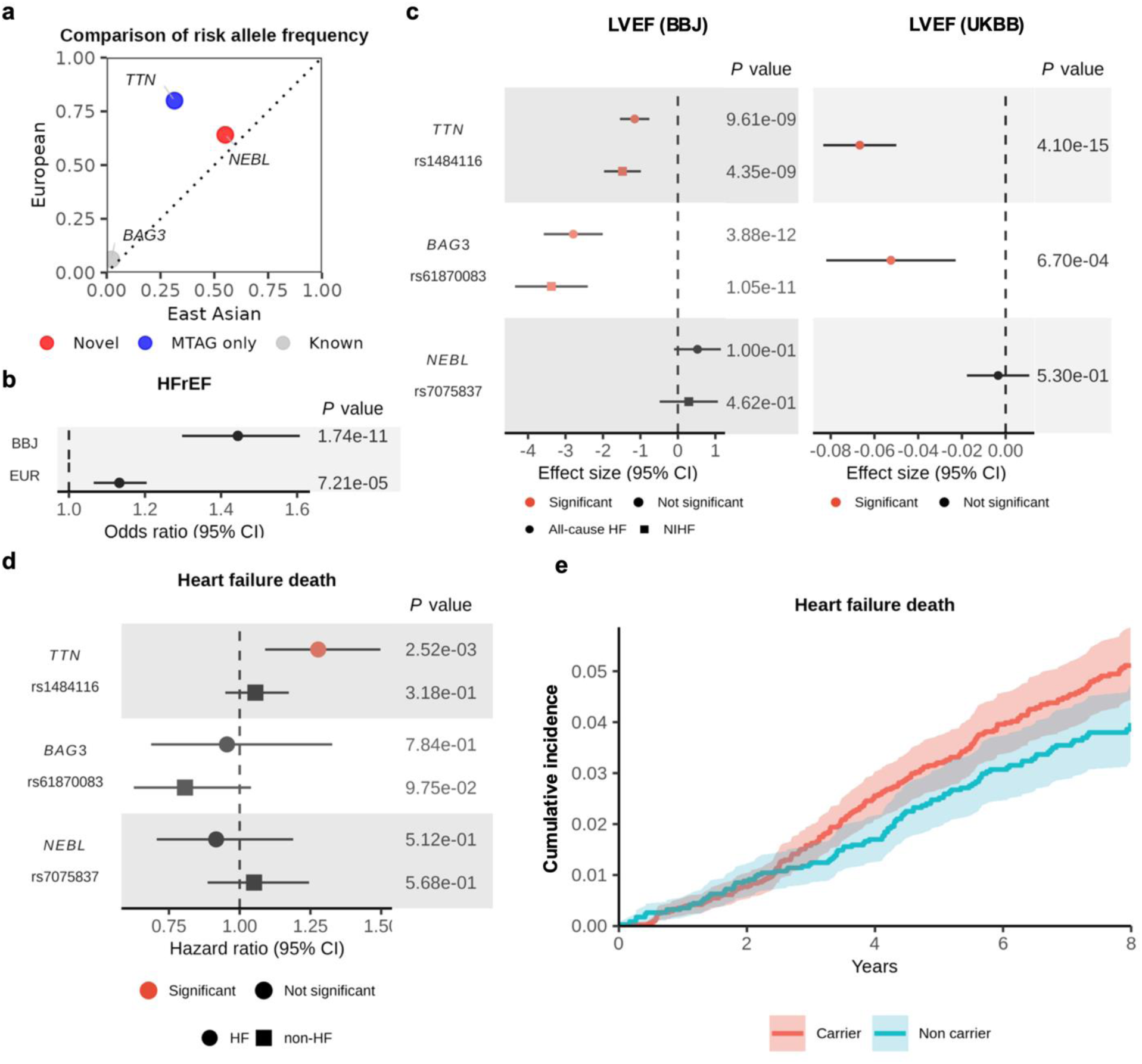
Effects of common variants in cardiomyopathy genes on left ventricular function and long-term HF mortality. **(a)** Minor allele frequency of lead SNPs in cardiomyopathy genes for East Asian and European populations according to gnomAD. **(b)** Effect size comparison of rs148116 (*TTN*) between BBJ and European for HFrEF. **(c)** Effects of lead variants in cardiomyopathy genes on LVEF in BBJ cohorts (left panel) and UKBB cohorts (right panel). Data are presented as estimated coefficients and their 95% confidence interval (CI). The shape of the points in c and d indicates the HF subtypes. Red points in c and d indicate statistically significant effects. HF, heart failure; NIHF, non-ischemic heart failure. **(d)** Effects of lead variants in cardiomyopathy genes for HF mortality among HF patients (indicated as circles, ICD-10 I50, 326 deaths among 8,481 individuals) and non-HF individuals (indicated as squares, ICD-10 I50, 749 deaths among 121,070 individuals). **(e)** Cumulative incidence curve for HF death according to carriers of rs1484116 (*TTN*) in HF patients.

### Development of a HF-PRS and its performance

Although most GWASs have been conducted in European populations, PRSs derived from European GWASs have shown poorer performance in other populations. On the other hand, PRSs derived from cross-ancestry GWASs have been reported to improve their accuracy in understudied populations^3^. Furthermore, integrating not only cross-ancestry GWAS of interest but also related phenotypes (e.g., blood pressure and LDL cholesterol for CAD) has proved to enhance the PRS performance^37^. Here, we split our case-control samples into the derivation dataset, dataset for linear combination, test dataset, and survival analysis dataset to avoid sample overlap (**Supplementary Fig. 14**). We derived each PRS from the Japanese all-cause HF, European/American/African all-cause HF, and European HFrEF, HFpEF, CAD, AF, and left ventricular mass with PRS-CS (see Methods section). The linear combination of these PRS was performed by Lasso regression with 10-fold cross-validation, resulting in the exclusion of PRSs calculated from Native American all-cause HF, and European NIHF. For the PRS derived from a single population GWAS, as concordant with the population specificity, the PRS derived from BBJ showed higher performance in the Japanese population than those from European (pseudo *R*^2^ = 0.709 in European versus 0.737 in BBJ; **Fig. 5a**) despite the smaller sample size. As suggested previously, cross-ancestry PRS showed higher performance than those from single population-derived PRS (**Fig. 5a** and **Supplementary Fig. 15a**). Additionally, combining HF-related phenotypes further improved the performance (**Fig. 5a** and **Supplementary Fig. 15a**). Hereafter, we used the best model (model J in **Fig. 5a**). The risk of developing HF in individuals with top PRS quintile was two times higher than that with bottom quintile (pseudo *R*^2^ with 95% CI for bottom quintile, 0.095 [0.084-0.107]; top quintile, 0.198 [0.183-0.214]; **Supplementary Fig. 15b**).

**Fig. 5.**
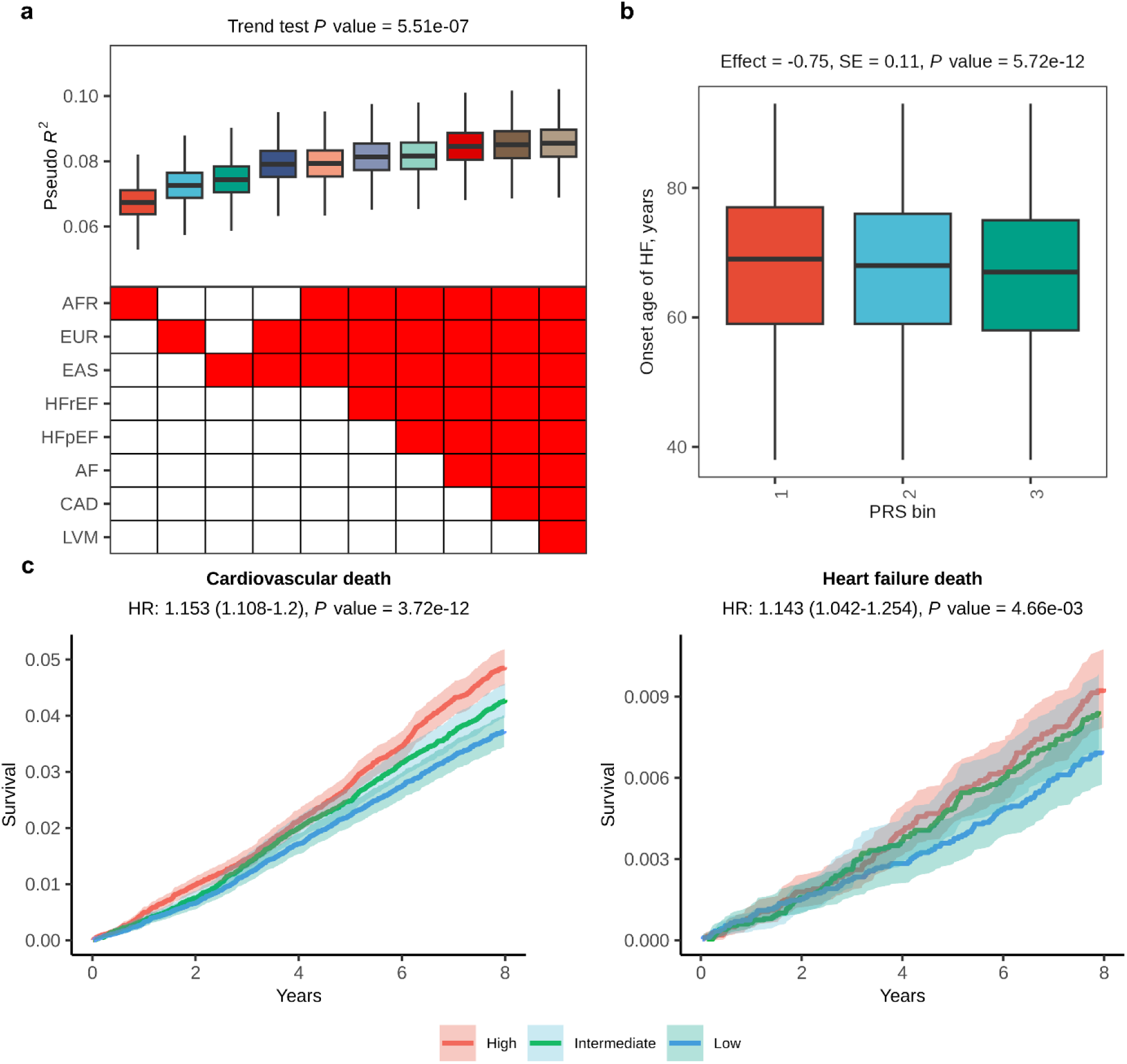
Performance and clinical impact of the HF-PRS. **(a)** Each point indicates Nagelkerke’s pseudo R^2^ for all-cause HF case-control status in the Japanese independent cohort (1,837 cases and 11,319 controls). PRS-derivation cohorts are indicated on the x-axis. **(b)** Association between HF-PRS and onset age of HF. The onset age of HF in individuals with data available (n = 10,810) is shown based on the HF-PRS tertiles. The number of individuals in each tertile is 3,603 to 3,604. The center line of the box plot indicates the median, the bounds represent the first and third quartile, and the whiskers reach to 1.5 times the interquartile range. **(c)** Kaplan-Meier estimates of cumulative events from cardiovascular mortality (left) and HF death (right) are shown with a band of 95%CI. Individuals are classified into high (top tertile), intermediate (middle tertile), and low PRS (bottom tertile). P values were calculated for PRS by Cox proportional hazard analysis and the significance was set at P = 8.3 × 10^−3^ (0.05/6).

### Impact of HF-PRS on HF phenotypes and outcomes

The impact of HF-PRS on long-term outcomes has not been previously evaluated. Earlier all-cause HF GWAS^2^ and HFrEF/HFpEF GWAS^5^ did not develop a PRS. Although the Global Biobank Meta-analysis Initiative (GBMI) developed an HF-PRS, it only assessed the predictive performance and not the clinical impact^38^. To assess the potential of the HF-PRS for clinical applications, we investigated the association between the PRS and the onset age of HF in individuals from our BBJ case samples (n =10,810). We observed that the onset age decreased as the PRS increased; individuals with the top tertile PRS were estimated to be approximately two years younger at HF onset compared to the bottom tertile individuals (**Fig. 5b**). To further explore the clinical utility of the HF-PRS, we assessed its impact on mortality using long-term follow-up data in BBJ. We stratified individuals without HF by tertiles (low, intermediate, and high groups) based on their PRS scores. The Kaplan–Meier estimates of cumulative cardiovascular mortality rate were significantly increased in the high PRS group. Furthermore, we demonstrated that the HF-PRS successfully stratified the risk of HF death (**Fig. 5c**).

## Discussion

We performed a large-scale GWAS with 16,251 HF cases and subtypes in the Japanese population along with cross-ancestry meta-GWAS and MTAG boosted by heart function-related traits and identified 19 novel loci. Additionally, through various gene prioritization approaches, we elucidated associations with clinical risk factors, cardiomyopathy, and drug targets. Furthermore, analysis of various datatypes in the biobank, including clinical and long-term prognosis data from biobanks, confirmed the identification of significant HF variants and the clinical utility of HF-PRS described here.

The Japanese GWAS identified 20 genome-wide significant loci associated with HF. This includes five new loci, where variants significantly prevalent in East Asians. We identified a novel association in the *CACNA1D* loci, suggesting the involvement of functional alterations in the calcium voltage-gated channel as a mechanism underlying HF. Since *CACNA1D* encodes the pore-forming α1 subunit of Ca_v_1.3 voltage-gated L-type calcium channels (LTCC) and is highly expressed in the sinus node and atrioventricular nodes^39^. Mutations in *CACNA1D* have been proved to cause sinoatrial node dysfunction^40^ suggesting the contribution of *CACNA1D* in the development of HF through chronotropic incompetence.

Furthermore, we performed cross-ancestry meta-analyses for HF subtypes, where 58 genome-wide significant loci were identified, resulting in the discovery of eight new loci. Among the prioritized genes based on the GWAS, *EDNRA,* and *CACNA1D* had a protective role against heart failure and are drug targets for other diseases. Both *EDNRA* inhibitors and *CACNA1D* inhibitors have already been reported to exacerbate HF, an observation that was successfully supported by our genetic analysis. We also identified a GIP/GLP-1-agonist as a drug repositioning candidate. By contrast, we genetically proved the detrimental effects of PDE3A and CDK4/6 blockade on HF pathogenesis.

In these analyses, we found that common variants of the cardiomyopathy genes *TTN*, *NEBL,* and *BAG3* contributed to the development of HF. MTAG additionally identified that common variants of *MYBPC3* were one of the susceptibility loci. Among these, a *TTN* common variant rs1484116 had a significant difference in the frequency and effect between Japanese and European populations. Notably, this common variant stratified individuals who went on to develop HF into those with a high mortality rate. Additionally, while rare truncating variants in the *TTN* gene comprise the most common genetic subtype of dilated cardiomyopathy, accounting for up to 25% of cases^41^, we show for the first time that the *TTN* common variant in a non-coding region also causes HF.

Integrating cross-ancestry GWAS of interest and GWAS of related phenotypes improves predictive performance in CAD^37^. In our HF-PRS, integrating HFrEF, HFpEF, AF, CAD, and left ventricular mass improved the performance. Moreover, we demonstrated that HF-PRS could predict HF death in subjects who had not yet been diagnosed as having HF.

Our study has several limitations. First, we did not have other large cohorts for the replication analysis. We did not replicate Japanese-specific HF loci using other East Asian cohorts since there were no other large East Asian biobanks available with enough of these patients. Instead, we assessed these variants with European data, showing the same effect direction. As for cross-ancestry meta-analyses, we did not have a non-overlapping corresponding cohort for all-cause HF and NIHF. Hence, we used European all-cause HF data for the replication of HFrEF and HFpEF cross-ancestry meta-analyses, showing almost the same effect direction. Second, we did not perform quantitative trait-based enrichment analysis (e.g. TWAS and splicing-TWAS) for the Japanese GWAS since there is no available molecular QTL dataset for the East Asian population. Thus, eQTL, sQTL, and other QTL datasets of the East Asian population are required in future research to further clarify the genetic differences between the East Asian and European populations.

In conclusion, our large-scale Japanese and cross-ancestry genetic analyses identified 19 new risk loci and provided insights into the distinct and shared genetic architecture of HF between Japanese and Europeans. Multiple prioritization methods allowed us to identify candidate genes involved in the pathogenesis. We also showed that the common variant of *TTN* can be a prognostic marker for HF patients in the Japanese population. Furthermore, analyses of disease prediction and long-term survival demonstrated the clinical utility of our HF-PRS. These data highlight the utility of HF genetics in clinical settings and provide useful evidence for the implementation of genomic medicine.

## Online Methods

### Study populations

We included two cohorts for the Japanese-specific meta-analysis including the BBJ 1^st^ cohort and BBJ 2^nd^ cohort from the BioBank Japan Project^42^. Further details on each cohort are described in the **Supplementary Note**.

For GWAS quality control, we excluded samples with a call rate < 0.98 and related individuals with pi-hat > 0.2, which is an index of relatedness based on pairwise identity-by descent implemented in PLINK 2.0 (20 Aug 2018 ver.). We then excluded samples with a heterozygosity rate > +4 s.d. We performed principal component analysis (PCA) using PLINK 2.0. and excluded outliers from the Japanese cluster. For the case samples in GWAS, we selected individuals with HF diagnosed by a physician based on accepted medical practice criteria. HFrEF was defined as having HF and LVEF < 40%, while HFpEF was defined as having HF and LVEF > 50%. NIHF was defined as having HF without CAD. The demographic features of the case-control cohort are shown in **Supplementary Table 1**.

### Genotyping, imputation, and quality control

To construct a population-specific reference panel, we included 7,825 Japanese WGS samples. Each sample was aligned to the human genome assembly GRCh38 and subjected to variant calling individually using the Illumina DRAGEN Bio-IT Platform (versions 3.4.12 and 3.5.7), generating gVCF files. Joint variant calling was subsequently performed with DRAGEN (version 3.6.3), employing default parameters and filtering criteria. Next variant quality control (QC) was conducted and variants from the jointly-called dataset were filtered out based on several exclusion criteria: 1) the presence of multi-alleles; 2) singleton; 3) missing data rate > 5%; 4) Hardy-Weinberg Equilibrium (HWE) p-value < 1×10^-6^. After QC, VCF files were phased on a whole-chromosome basis using SHAPEIT4 (version 4.2)^43^.

GWAS subjects were genotyped using the Illumina Human OmniExpress Genotyping BeadChip or a combination of Illumina HumanOmniExpress and HumanExome BeadChips (Illumina, San Diego, CA). For genotype quality control (QC), we excluded variants with (i) SNP call rate < 0.99, (ii) MAF < 0.01, and (iii) Hardy–Weinberg equilibrium P value < 1.0 × 10^−6^. BBJ 1^st^ and BBJ 2^nd^ cohorts were separately phased using SHAPEIT2 (version r837) and EAGLE2 (version 2.39), respectively. Imputation was then carried out with Minimac4 (version 1.0.0)^44^ with a minor adjustment: variant positions in the array data, initially based on hg19, were remapped to GRCh38 using BLAST based on the sequence of flanking sites.

### GWAS

In the Japanese GWAS, association was performed by logistic regression analysis assuming an additive model with adjustment for age, age^2^, sex, and top 10 principal components (PCs) using REGENIE^45^. We selected variants with Minimac4 imputation quality score of > 0.3 and MAF ≥ 0.01. The genome-wide significance threshold was defined at *P* < 5.0 × 10^-8^. The genomic inflation factor (λGC) was 1.044, 1.080, 1.050, and 1.083 for all-cause HF, HFrEF, HFpEF, and NIHF, respectively. (**Supplementary Figure. 2a**). Adjacent genome-wide significant SNPs were grouped into one locus if they were within 1 Mb of each other. We defined a locus as follows: (1) extracted genome-wide significant variants (*P* < 5.0 × 10^-8^) from the association result, (2) added 500 kb to both sides of these variants, and (3) merged overlapping regions. If the locus did not contain coordinates with previously reported genome-wide significant variants (i.e., all variants with *P* < 5.0 × 10^-8^ in the previously reported locus), the region was annotated as being novel. To identify independent association signals in the loci, we conducted a stepwise conditional analysis for genome-wide significant loci defined as described in the GWAS. First, we performed logistic regression conditioning on the lead variants of each locus. We set a locus-wide significance at *P* < 5.0 × 10^-6^ and repeated this procedure until none of the variants reached locus-wide significance for each locus.

### Linkage-disequilibrium score regression and heritability

To estimate confounding biases such as cryptic relatedness and population stratification, we performed a linkage-disequilibrium score regression (LDSC) (Version 1.0.0). We selected SNPs with MAF ≥ 0.01 and variants within the major histocompatibility complex region were excluded. For the regression, we used the East Asian LD scores provided by the authors (https://github.com/bulik/ldsc/). The all-cause HF population prevalence was set at 6.5% according to the Japan Medical Data Center and the Medical Data Vision datasets^46^. Since 37.4%, 45.1%, and 26.6% of all-cause HF were HFrEF, HFpEF, and NIHF, we assumed population prevalence was 2.4% for HFrEF, 2.9% for HFpEF, and 1.7% for NIHF^47^.

### Cross-ancestry meta-analysis

All genomic coordinates were converted to GRCh37 with the liftover tool. Fixed-effect meta-analyses based on inverse-variance weighting were performed for all HF phenotypes. We defined genome-wide significant loci by iteratively spanning the ±500 kb region around the most significant variant and merging overlapping regions until no genome-wide significant variants were detected within ±1 Mb. A locus was categorized as “known” if the region after merging was within ±500kb of variants for the corresponding phenotype in previous all-cause HF, HFrEF, HFpEF, NIHF GWAS, “previously reported only in MTAG” if in previous all-cause HF MTAG results^2^, otherwise, it was categorized as a novel. The most significant variant in each locus was selected as the index variant. Cochran’s Q-test for heterogeneity was conducted to identify loci with index variants that have different effect sizes across GWAS data sets. We filtered variants with strong heterogeneity (*P_het_* < 1.0 × 10^-4^) or MAF < 1%.

### Multi-trait association study

To identify additional HF-risk loci, we used MTAG^20^ in Europeans, including cardiac MRI traits, which were most strongly correlated with HF. We included left ventricular mass as it was most strongly correlated with all HF phenotypes (**Supplementary Figure 9a)**. We filtered variants with MAF < 1% and defined genome-wide significant loci in the same way as trans-ancestry meta-analysis.

### Independent follow-up of GWAS signals

We sought to replicate internally the HF-risk loci reaching genome-wide significance in the Japanese GWAS meta-analyses within BBJ1 and BBJ2. As for genome-wide significant variants identified in the cross-ancestry meta-analysis, we assessed EUR and EAS. As external replication of associations, we assessed significant variants identified in HFrEF or HFpEF using European all-cause HF.

### Genetic correlation of HF and cardiac imaging traits

Cross-trait LDSC was performed to estimate genetic correlation (r_g_) between HF and cardiac MRI traits. LDSC is a computationally efficient method that utilizes GWAS summary statistics to estimate heritability and genetic correlation between polygenic traits while accounting for sample overlap.

### Transcriptome-wide association study

We performed a transcriptome-wide association study (TWAS) to assess gene-HF associations using FUSION^48^, which estimates the association between predicted gene expression levels and a phenotype of interest using summary statistics and gene expression prediction models. We used precomputed prediction models of gene expression in tissues considered to be relevant for HF with LD reference data in GTEx v8 and the summary statistics of the trans-ancestry HF-GWAS as input. Furthermore, we used the cross-tissue weights generated in GTEx v8 using sparse canonical correlation analysis (sCCA) across 49 tissues available on the FUSION website, including gene expression models for the first three canonical vectors (sCCA1-3), which were shown to capture most of the gene expression signal.

Transcriptome-wide significant genes (eGenes) and the corresponding eQTLs were determined using the Bonferroni correction, based on the average number of features (8,036.5 genes) tested across all reference panels and correcting for the four HF phenotypes (*P* < 1.55 × 10^−6^). To ensure that the observed associations did not reflect a random correlation between gene expression and non-causal variants associated with stroke, we performed a colocalization analysis on the eGenes to estimate the posterior probability of a shared causal variant between the gene expression and trait association (PP4). eGenes presenting a PP4 ≥ 0.75 were considered to be significant.

### Splicing-TWAS

We performed a splicing-TWAS to assess splicing-HF associations using MetaXcan. We used GTEx v8 MASHR-based models (https://github.com/hakyimlab/MetaXcan). The significant splicing sites were determined using the Bonferroni correction, based on the number of splicing sites multiplied by the four HF phenotypes.

### Identifying protein-altering variants

To identify protein-altering variants among our genome-wide significant associations, we took the sentinel variants and their LD proxies at r^2^ ≥ 0.8 as estimated in the European ancestry subset of 1000 Genomes Project phase 3 and annotated them using the Ensembl VEP. We selected for each sentinel variant any proxies identified as having a ‘high’ (that is, stop-gain and stop-loss, frameshift insertion and deletion, donor and acceptor splice-site and initiator codon variants) or ‘moderate’ (that is, missense, in-frame insertion and deletion, splice region) consequence and recorded the gene that the variant disrupts.

### Polygenic prioritization of candidate causal genes

We implemented PoPS, a similarity-based gene prioritization method designed to leverage the full genome-wide signal to nominate causal genes independent of methods utilizing GWAS data proximal to the gene (https://github.com/FinucaneLab/pops.git). Broadly, PoPS leverages polygenic enrichments of gene features including cell-type-specific gene expression, curated biological pathways, and protein-protein interaction networks to train a linear model to compute a PoPS for each gene.

### Variants responsible for cardiovascular-relevant monogenic disorders

To identify genes harboring pathogenic variants responsible for HF-relevant monogenic disorders, we searched the NCBI’s ClinVar database (https://www.ncbi.nlm.nih.gov/clinvar/). Variants were pruned to those within ±500 kb of our HF sentinel variants; categorized as ‘pathogenic’ or ‘likely pathogenic’; with a listed phenotype; and with either (i) details of the evidence for pathogenicity, (ii) expert review of the gene or (iii) a gene that appears in practice guidelines^49^. We then filtered the variants that were annotated with a manually curated set of cardiovascular-relevant phenotype terms, including those related to HF and HF risk factors (**Supplementary Tables 12**, **24**, and **35**). Where a variant was annotated with multiple genes, both genes were considered as potentially pathogenic.

### Phenotyping knock-out mice

Human gene symbols were mapped to gene identifiers (HGNC) and mouse ortholog genes were obtained using Ensembl (www.ensembl.org). Phenotype data for single-gene knock-out models were obtained from the International Mouse Phenotyping Consortium, data release 19.1 (www.mousephenotype.org), and from the Mouse Genome Informatics database, data from July 2023 (www.informatics.jax.org). For each mouse model, reported phenotypes were grouped using the mammalian phenotype ontology hierarchy into broad categories relevant to HF as follows: abnormal glucose homeostasis (MP:0002078), abnormal blood coagulation (MP:0002551), muscle phenotype (MP:0005369), abnormal lipoprotein level (MP:0010329), abnormal catecholamine level (MP:0011479), abnormal myofibroblast differentiation (MP:0012196), cardiac edema (MP:0012270), abnormal lipid metabolism (MP:0013245), adipose tissue necrosis (MP:0013249), abnormal susceptibility to non-insulin-dependent diabetes (MP:0020086), abnormal fibroblast physiology (MP:0020414), abnormal adipose tissue noradrenaline turnover (MP:0021021), cardiac amyloidosis (MP:0021148) along with phenotypes previously used in CAD analysis^49^ (**Supplementary Table 46**). This resulted in mapping from genes to phenotypes in animals (**Supplementary Tables 14, 25,** and **36**).

### PRS derivation and performance

First, we divided our dataset into three groups: (i) a discovery group to derive PRS (12,335 cases and 107,333 controls), (ii) a group used for linear combination (1,837 cases and 11,320 controls), (iii) a test group to assess PRS performance (1,837 cases and 11,319 controls), and (iv) a group for the survival analysis (67,991 controls) (**Supplementary Fig. 14**). Next, we used PRS-CS to derive each PRS from Japanese-, European-, American-, African-all-cause HF, European HFrEF, HFpEF, NIHF, AF, CAD, and left ventricular mass. We used LD reference panels constructed using the 1000 Genomes Project phase 3 samples according to each cohort population. Subsequently, we calculated PRSs in the withheld BBJ cohorts for linear combination and conducted logistic regression with L1 regularization and 10-fold cross-validation. Finally, we used the Japanese all-cause HF, European all-cause HF, European HFrEF, European HFpEF, European CAD, European AF, and European left ventricular mass to construct the HF-PRS. The performance of the PRS was measured as (1) Nagelkerke’s pseudo-*R*^2^ obtained by modeling age, sex, the top 10 PCs, and normalized PRS and (2) the area under the curve of the receiver operator curve in the same model as Nagelkerke’s pseudo *R*^2^. Using the models previously described, we calculated the PRSs and assessed their performance for the independent test cohort.

### Association of HF-PRS with age of HF onset

To assess the association between HF-PRS and age at HF onset, we extracted HF case samples with available data on age at HF onset (n = 10,810, the median age of HF onset was 68 years of age [IQR 60–76]) and constructed a linear regression model of age at HF onset including HF PRS along with sex and the top 10 PCs to estimate effects of HF-PRS on age of AF onset.

### Survival analysis

For survival analysis, the Cox proportional hazards model was used to assess the association between HF-PRS and long-term mortality. We obtained survival follow-up data with the ICD-10 code for 132,737 individuals from the BBJ dataset^42^. The causes of death were classified into three categories according to ICD-10 codes: cardiovascular death (I00–I99) and HF death (I50). For HF-PRS analysis, we used a holdout survival analysis cohort (**Supplementary Figure 10**). The median follow-up period was 8.4 years (IQR 6.8 – 9.9). The Cox proportional hazards model was adjusted for sex, age, the top 10 PCs, and disease status.

Analyses were performed with the R package survival v.2.44, and survival curves were estimated using the R package survminer v.0.4.6, with modifications.

## Data availability

Summary statistics of Japanese GWAS and polygenic risk scores derived in this study will be publicly available at the National Bioscience Database Center (https://humandbs.dbcls.jp/en/). The GWAS summary statistics for HF (GBMI American: https://www.globalbiobankmeta.org/; Kadoorie: https://www.ckbiobank.org/; FinnGen NIHF: https://console.cloud.google.com/storage/browser/_details/finngen-public-data-r9/summary_stats/, R9_I9_NONISCHCARDMYOP; UKBB NIHF: https://cvd.hugeamp.org/), and cardiac MRI (left ventricle, right ventricle and atrium: http://kp4cd.org/datasets/mi; left atrium: https://zenodo.org/records/5074929; left ventricular mass: https://cvd.hugeamp.org/) traits are publicly available.

## Code Availability

The code used in this study is available on reasonable request to the corresponding authors.

## Acknowledgements

We thank the staff of BBJ for their assistance in collecting samples and clinical information. We thank Naoko Miyagawa, Akio Wada, and Shuji Ito for their technical assistance. This research was funded by the Japan Agency for Medical Research and Development (AMED) (JP24bm1423005 to K.I, nos. JP24ek0210164, JP24tm0524004, and JP24tm0624002 to K.I., S.N., and I.K., and nos. JP24km0405209 and JP20ek0109487 to K. Miyazawa, K. I, S. K., S. N., H. Akazawa, and I. K.), Japan Foundation for Applied Enzymology (to N.E), the Japan Society for the Promotion of Science (to N. E. and K. I.), and the Research Funding for Longevity Sciences from NCGG (24–15 to K.O. and K.I.). BBJ was supported by the Tailor-Made Medical Treatment Program of the Ministry of Education, Culture, Sports, Science, and Technology (MEXT) and AMED under grant numbers JP17km0305002, JP17km0305001, and JP24tm0624002.

## Author Contributions

N. E., K. I., C. T., Y. K., T. N., and I. K. conceived and designed the study. K. I, C. T., K. Matsuda., Y. M., and Y. K. collected and managed the BBJ sample. T. N, Y. M., and Y. K. performed the genotyping. N. E, K. Miyazawa, S. K, H. I., R.K., H.Y., F.T., M.F., R.O. performed the statistical analyses. N.E, K. I, C. T, S. K., K.T., X.L, K.O., Y. O., T.Y., Y. K., T.N., K.N., H. Akazawa, P.T.E., MG.L., SM.D., BF.V, J.J., and YV.S. contributed to data collection, processing, analysis, and interpretation. K. I, C.T., Y K., H. Aburatani, P.T.E. and I.K. supervised the study. N.E. and K.I. wrote the manuscript, and many authors have provided valuable edits.

## Supplementary Notes

### 1. The SNP heritability and the liability-scale heritability

The proportion of the variation in HF phenotypes (the single nucleotide polymorphism (SNP) heritability; h^2^) was estimated to be 1.5% (standard error of the mean [s.e.m.] 0.3%) for all-cause HF, 1.5% (0.3%) for HFrEF, 0.5% (0.3%) for HFpEF, and 1.1% (0.3%) for NIHF using linkage disequilibrium (LD)-score regression (LDSC). The liability-scale h^2^ was estimated at 4.8% (s.e.m. 1.0%) for all-cause HF, 12.6% (2.9%) for HFrEF, 2.8% (1.7%) for HFpEF, and 3.1% (1.0%) for NIHF, suggesting the heterogeneous heritability in each HF phenotype.

### 2. Replication of previously identified HF risk loci

We assessed the 47 independent variants previously reported from all-cause HF GWAS^1^, 13 variants from HFrEF GWAS, one variant from HFpEF GWAS^2^, and one variant from NIHF^3^ in our Japanese GWAS (BBJ). Out of 47 variants identified in the previous all-cause HF GWAS, 34 variants or variants tagged with the lead variant existed in BBJ, 28 had the same effect direction, and 18 were nominally significant (*P* < 0.05; **Supplementary Fig. 2, Supplementary Table 3**). Out of 13 variants identified in the European HFrEF GWAS, 11 existed in BBJ, 10 had the same effect direction, and 3 were nominally significant. The lead variant identified in the European HFpEF GWAS had the same direction and was nominally significant in BBJ, and the lead or tagged variant identified in the European NIHF GWAS did not exist in BBJ.

### 3. Internal and external replication for novel loci found in BBJ GWAS

We sought to replicate the HF-risk loci that reached genome-wide significance in individual GWAS cohorts. The effect size, direction (**Supplementary Fig. 3a**), and allele frequency of the identified lead variants (**Supplementary Figure 3b**) were strongly concordant in BBJ 1^st^ and BBJ 2^nd^ cohorts for all HF phenotypes (Pearson’s correlation: all-cause HF 0.981; HFrEF 0.961; HFpEF 0.752; NIHF 0.992). Three out of the five newly identified loci were nominally significant in both BBJ 1^st^ and BBJ 2^nd^ cohorts (**Supplementary Fig. 3a** and **Supplementary Table 4**). Out of the five newly identified loci, three loci identified in all-cause HF were found in the European GWAS. rs35593046 were successfully replicated with nominal association (*P* < 0.05) with the same effect direction (**Supplementary Fig. 3c** and **Supplementary Table 5**). As for rs6471480 and rs7075837, the same effect direction was observed in the European population. rs76704104 identified in HFrEF and rs78228190 identified in HFpEF were not observed in the European population according to the Genome Aggregation Database v4.0.0 (gnomAD) (https://gnomad.broadinstitute.org/).

### 4. Cohorts used for cross-ancestry meta-analyses

Cross-ancestry datasets (**Supplementary Table 18**) yielded 120,203 cases for all-cause HF (BBJ: 16,251, EUR: 95,524, KADOORIE: 1,467, AMR: 1,170, AFR: 5,791), 23,749 cases for HFrEF (BBJ 4,254, EUR: 19,495), 26,743 cases for HFpEF (BBJ: 7,154, EUR: 19,589), and 22,864 cases for NIHF (BBJ: 11,122, UKBB: 1,816, FinnGen: 9,926). The number of control samples was 1,502,645 (BBJ: 197,577, EUR: 1,270,968, KADOORIE: 75,149, AMR: 13,217, AFR: 20,883) for all-cause HF, 456,520 for HFrEF/HFpEF (BBJ: 197,577, EUR: 258,943), and 863,254 for NIHF (BBJ: 171,995, UKBB: 387,652, FinnGen: 303,607), respectively. We included variants with MAF ≥ 1% and tested a total of 11,119,536 variants for all-cause HF, 4,800,324 variants for HFrEF, 4,807,066 variants for HFpEF, and 4,998,828 variants for NIHF.

### 5. Internal and external replication of cross-ancestry meta-analyses results

The effect size and direction of the identified lead variants (**Supplementary Fig. 7b**) were concordant in East Asians and Europeans for all HF phenotypes (Pearson’s correlation: all-cause HF 0.898; HFrEF 0.864; NIHF 0.983) except for HFpEF, for which Pearson’s correlation could not be calculated with significant loci < 3.

To replicate lead variants of HFrEF and HFpEF using independent datasets, these were assessed with European all-cause HF GWAS since there were no other available HFrEF/HFpEF GWAS datasets. As for all-cause HF and NIHF, there was no dataset for external replication. Effect sizes were generally concordant with Pearson’s correlation of 0.817 in HFrEF (**Supplementary Figure 7d** and e). The effect direction was also concordant in all of the three novel loci (**Supplementary Table 21**). As for HFpEF, both significant variants were known HF-related variants.

### 6. Prioritization of rs10851802 in the cross-ancestry meta-analyses

Among novel loci, we could not prioritize a gene for rs10851802. rs10851802 is linked to *GLCE* according to the Open Targets database. rs10851802 was associated with rs2415040 (r^2^ = 0.548), which was a variant for lean mass^4^. Lean mass is associated with incident HF^5^. Although we could not prioritize a gene in this locus, this evidence suggests that this locus was one of the disease susceptibility loci. Further investigations were warranted to identify the candidate gene in this locus.

### 7. Prioritized genes in MTAG

*BDNF* prioritized by PoPS (**Supplementary Table 40**) harbored pathogenic variants responsible for obesity (**Supplementary Table 38**), and its knockout showed the same phenotype (**Supplementary Table 39**). *MYBPC3* was one of the known cardiomyopathy genes (**Supplementary Table 14**, **38** and **39**), and this is the first report to show that common variants of *MYBPC3* were associated with HF. *SLC4A7*, prioritized by PoPS (**Supplementary Table 40**), was associated with abnormal vasoconstriction (**Supplementary Table 39**). Considering *SLC4A7* splicing was enriched in fibroblasts (**Supplementary Figure 10b**), it suggested that arterial fibroblasts contributed to HF through hypertension. Common variants in *PCSK1* influenced blood pressure^6^, which was consistent with the fact that *PCSK1* was prioritized H-MAGMA using the aorta Hi-C dataset (**Supplementary Fig. 10c**) and *PCSK1* knockout showed obesity and hypertension (**Supplementary Table 39**). *PTPRJ* knockout showed abnormal vascular development and abnormal heart development (**Supplementary Table 39**). *PLPP3* was enriched in the fibroblasts in TWAS (**Figure 10a**), and its knockout caused the abnormal vasculogenesis. These results were consistent with the previous report which indicated that *PLPP3* regions was associated with hypertension^7^ and coronary artery disease^8^.

Among the novel loci, we could not assign a gene to rs10888955. This variant was strongly associated with reticulocyte count^9^. Given that previous Mendelian randomization analysis revealed the positive association of CAD with reticulocytes^10^, this locus could contribute to HF development through CAD.

## Supplementary methods

### Cohort characteristics for BBJ 1^st^ cohort and BBJ 2^nd^ cohort

All subjects were Japanese and were registered in the BioBank Japan (BBJ) project (https://biobankjp.org/). The BBJ is a hospital-based national biobank project that collects DNA and serum samples and clinical information from 12 cooperative medical institutes throughout Japan (Osaka Medical Center for Cancer and Cardiovascular Diseases, Cancer Institute Hospital of Japanese Foundation for Cancer Research, Juntendo University, Tokyo Metropolitan Geriatric Hospital, Nippon Medical School, Nihon University School of Medicine, Iwate Medical University, Tokushukai Hospitals, Shiga University of Medical Science, Fukujuji Hospital, National Hospital Organization Osaka National Hospital, and Iizuka Hospital). Approximately 267,000 patients with any of the 47 target diseases were enrolled from 2003 to 2007, and from 2013 to 2018. All subjects were at least 18 years old. Informed consent was obtained from all participants, and our study was approved by the relevant ethical committee at each facility.

### Cell type-specific heritability enrichment of disease associations using stratified LD score regression

We applied stratified LD score regression (S-LDSC)^11^ (v1.0.1. https://github.com/bulik/ldsc) to the GWAS summary statistics of the cross-ancestry meta-analysis to evaluate the contribution of genetic variation in cell type-specific genes to trait heritability. Heritability enrichment per cell type was considered significant at *P* < 7.2 × 10^-5^ (0.05/694) based on the number of cell types tested (694).

### Links between medication and prioritized loci

We searched medications linked to prioritized genes using DrugBank and the Therapeutic Target Database. Among medications with the same pharmacological actions, we chose a representative one manually. The role of prioritized genes (e.g. protective or detrimental against HF) was determined based on (1) knockout mice phenotyping or (2) the direction of TWAS Z-score if knockout mice phenotyping was not reported to be associated with HF.

### miRNA enrichment analysis

The overall polygenic contribution of miRNA–target gene network to the traits through the tissue-naïve approach was estimated using MIGWAS software with default settings^12^. The significance threshold was set at 0.0125 based on the number of HF phenotypes (0.05/4). Candidate miRNA-gene pairs were estimated by MIGWAS with a significance threshold set at FDR < 0.05 for miRNA and genes.

### Heritability enrichment with scDRS

To determine which cell types in hearts were enriched for HF GWAS, we used scDRS (v.1.0.2)^13^ with default settings. We used MAGMA v.1.10^14^ to map single-nucleotide polymorphisms to genes (GRCh37 genome build from the 1000 Genomes Project) using an annotation window of 0 kb. We used the resulting annotations and GWAS summary statistics to calculate each gene’s MAGMA z score (association with a given trait). The 1,000 disease genes used for scDRS were chosen and weighted based on their top MAGMA z scores. To determine trait association at the annotated cell type resolution, we used the z scores computed from scDRS’s downstream Monte Carlo test. These Monte Carlo z scores were converted to theoretical P values using a one-sided test under a normal distribution. The significance threshold was set at *P* < 2.4 × 10^-3^ based on Bonferroni correction (0.05/21 based on the number of cell types).

### Members of participating consortia The Biobank Japan Project

Koichi Matsuda^1,2^, Takayuki Morisaki^2,3^, Yukinori Okada^4^, Yoichiro Kamatani^5^, Kaori Muto^6^, Akiko Nagai^6^, Yoji Sagiya^2^, Natsuhiko Kumasaka^7^, Yoichi Furukawa^8^, Yuji Yamanashi^3^, Yoshinori Murakami^3^, Yusuke Nakamura^3^, Wataru Obara^9^, Ken Yamaji^10^, Kazuhisa Takahashi^11^, Satoshi Asai^12,13^, Yasuo Takahashi^13^, Shinichi Higashiue^14^, Shuzo Kobayashi^14^, Hiroki Yamaguchi^15^, Yasunobu Nagata^15^, Satoshi Wakita^15^, Chikako Nito^16^, Yu-ki Iwasaki^17^, Shigeo Murayama^18^, Kozo Yoshimori^19^, Yoshio Miki^20^, Daisuke Obata^21^, Masahiko Higashiyama^22^, Akihide Masumoto^23^, Yoshinobu Koga^23^ & Yukihiro Koretsune^24^

^1^Laboratory of Genome Technology, Human Genome Center, Institute of Medical Science, The University of Tokyo, Tokyo, Japan.

^2^Laboratory of Clinical Genome Sequencing, Graduate School of Frontier Sciences, The University of Tokyo, Tokyo, Japan.

^3^The Institute of Medical Science, The University of Tokyo, Tokyo, Japan.

^4^Department of Genome Informatics, Graduate School of Medicine, The University of Tokyo, Tokyo, Japan.

^5^Laboratory of Complex Trait Genomics, Graduate School of Frontier Sciences, The University of Tokyo, Tokyo, Japan.

^6^Department of Public Policy, Institute of Medical Science, The University of Tokyo, Tokyo, Japan.

^7^Division of Digital Genomics, Institute of Medical Science, The University of Tokyo, Tokyo, Japan.

^8^Division of Clinical Genome Research, Institute of Medical Science, The University of Tokyo, Tokyo, Japan.

^9^Department of Urology, Iwate Medical University, Iwate, Japan.

^10^Department of Internal Medicine and Rheumatology, Juntendo University Graduate School of Medicine, Tokyo, Japan.

^11^Department of Respiratory Medicine, Juntendo University Graduate School of Medicine, Tokyo, Japan.

^12^Division of Pharmacology, Department of Biomedical Science, Nihon University School of Medicine, Tokyo, Japan.

^13^Division of Genomic Epidemiology and Clinical Trials, Clinical Trials Research Center, Nihon University. School of Medicine, Tokyo, Japan.

^14^Tokushukai Group, Tokyo, Japan.

^15^Department of Hematology, Nippon Medical School, Tokyo, Japan.

^16^Laboratory for Clinical Research, Collaborative Research Center, Nippon Medical School, Tokyo, Japan.

^17^Department of Cardiovascular Medicine, Nippon Medical School, Tokyo, Japan.

^18^Tokyo Metropolitan Geriatric Hospital and Institute of Gerontology, Tokyo, Japan.

^19^Fukujuji Hospital, Japan Anti-Tuberculosis Association, Tokyo, Japan.

^20^The Cancer Institute Hospital of the Japanese Foundation for Cancer Research, Tokyo, Japan.

^21^Center for Clinical Research and Advanced Medicine, Shiga University of Medical Science, Shiga, Japan.

^22^Department of General Thoracic Surgery, Osaka International Cancer Institute, Osaka, Japan.

^23^Iizuka Hospital, Fukuoka, Japan.

^24^National Hospital Organization Osaka National Hospital, Osaka, Japan.

---

**Supplementary Fig. 1.**
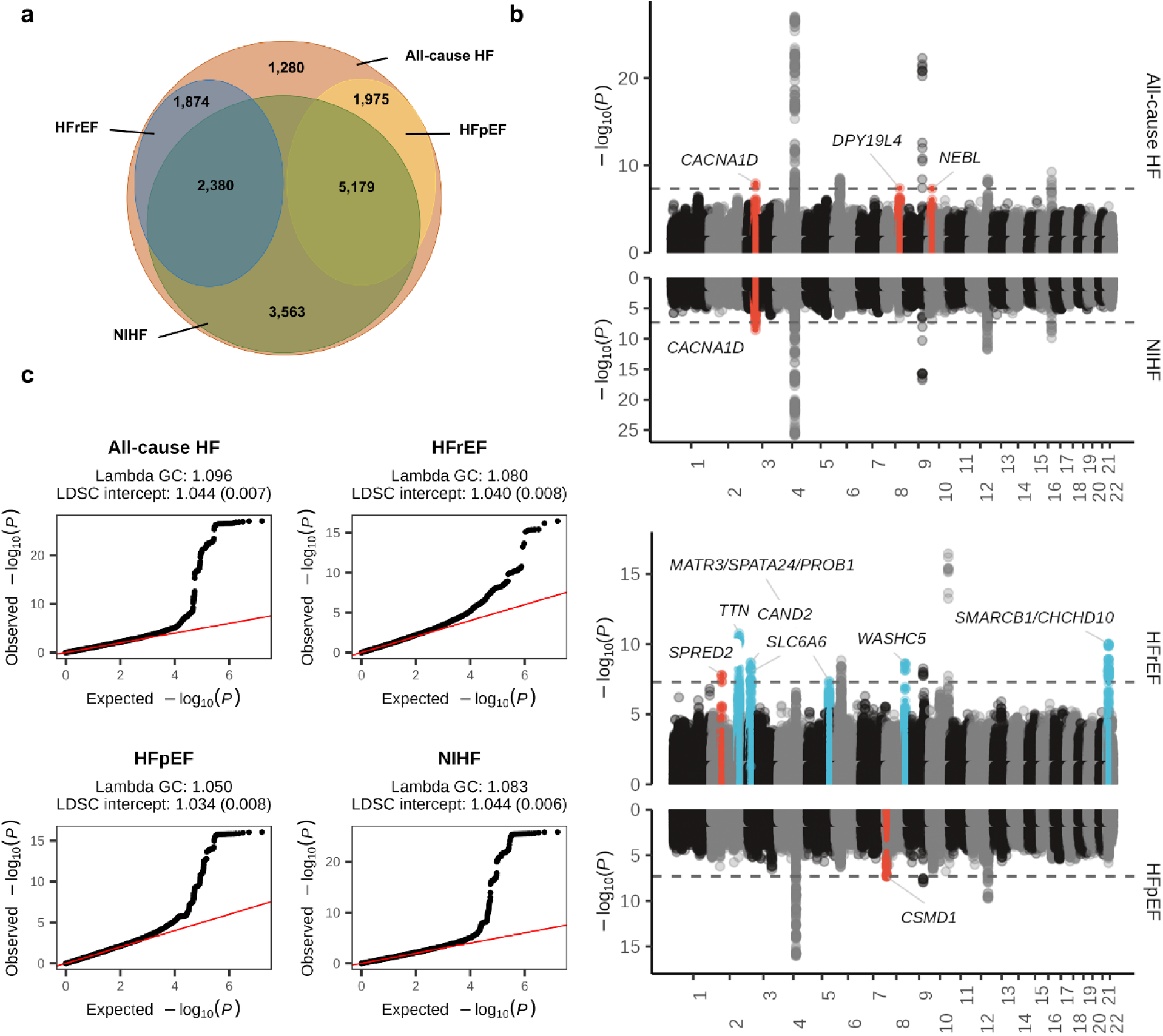
Results of Japanese GWAS. **(a)** Sample overlap between each HF phenotype in Japanese GWAS. **(b)** Miami plot for four HF phenotypes. -log_10_(*P* value) on the *y*-axis are shown against the genomic positions (hg19) on the *x*-axis. Association signals that reached a genome-wide significance level (P < 5 × 10^-8^) are shown in red if new loci and in blue if previously reported only in MTAG but not in GWAS. **(c)** QQ plot for each HF phenotype in Japanese GWAS. Two-sided *P* values were calculated using a logistic regression model. GWAS, genome-wide association study; HF, heart failure; MTAG, multi-trait analysis of GWAS.

**Supplementary Fig. 2.**
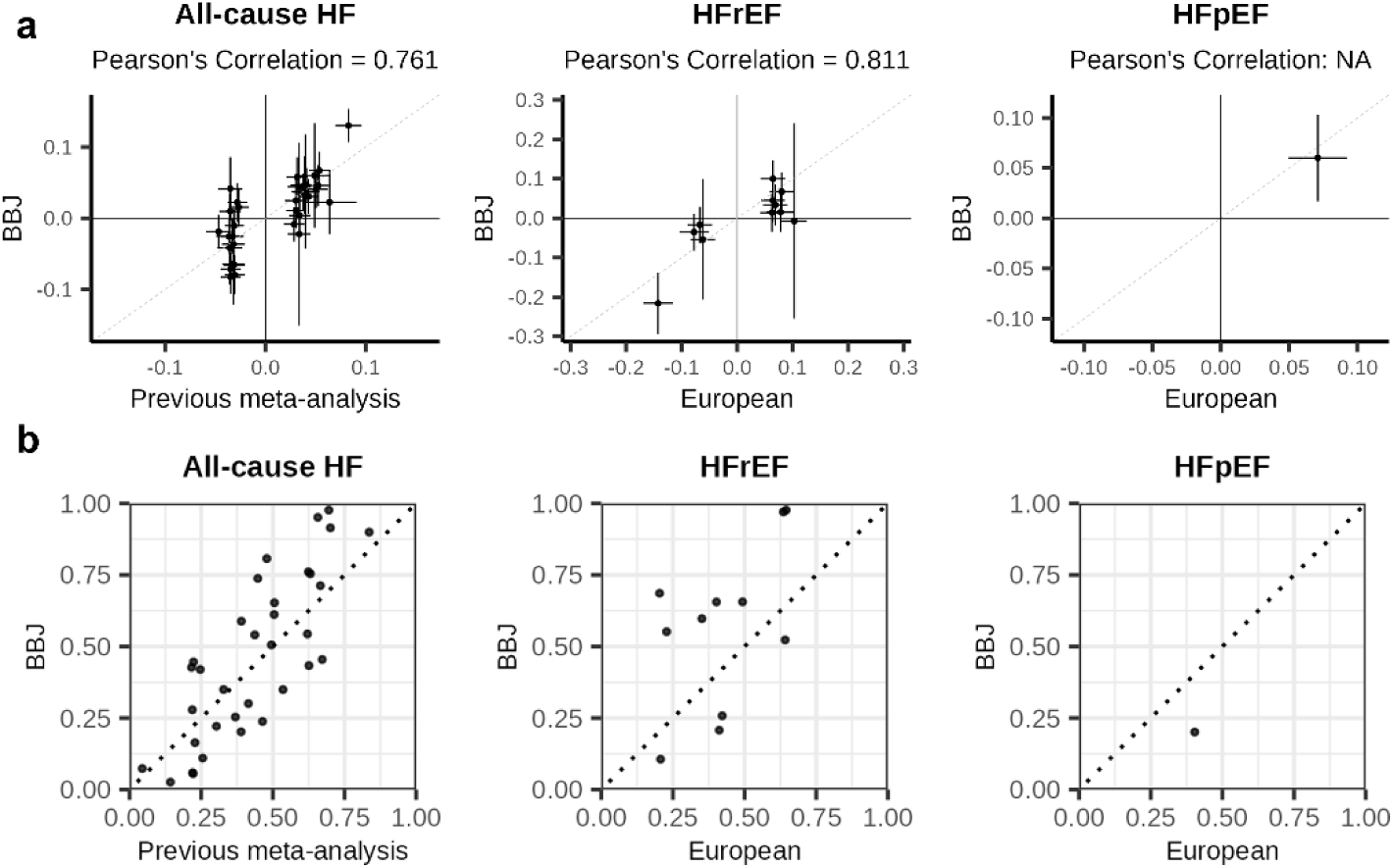
Validation of previously identified HF loci in BBJ. Comparison of allelic effects **(a)** and allele frequency **(b)** between the previous GWAS and BBJ. Effect size with its 95%CI or allele frequency of previous GWAS are shown in the *x*-axis, and those of BBJ are shown in the *y-*axis.

**Supplementary Fig. 3.**
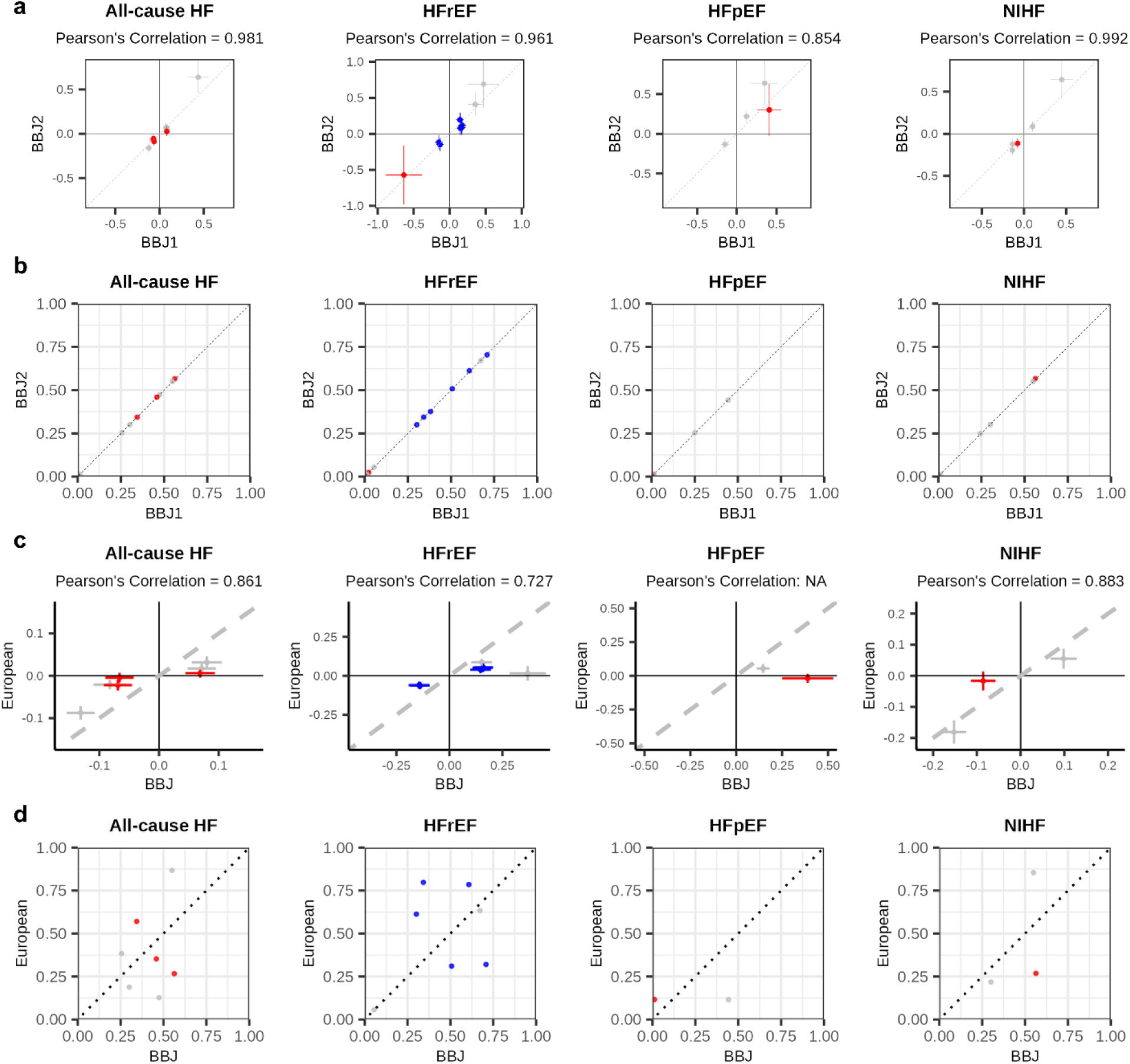
Comparison of allele frequencies and allelic effects identified in Japanese GWAS. (**a** and **b**) Comparison of allelic effects **(a)** and allele frequency **(b)** between BBJ 1^st^ and BBJ 2^nd^. Effect size with its 95%CI or allele frequency of BBJ 1^st^ GWAS are shown in the *x*-axis, and those of BBJ 2^nd^ are shown in the *y*-axis. **(c** and **d)** Comparison of allelic effects (**c**) and allele frequencies (**d**) between our Japanese GWAS (BBJ) and European GWAS. Effect size and its 95%CI, or allele frequencies of BBJ are shown in the *x*-axis, and those of European GWAS are shown in the *y*-axis. Points are shown in red if new loci and in blue if previously reported only in MTAG but not in GWAS. CI, confidence interval; GWAS, genome-wide association study; HF, heart failure; MTAG, multi-trait analysis of GWAS; QQ, quantile-quantile.

**Supplementary Fig. 4.**
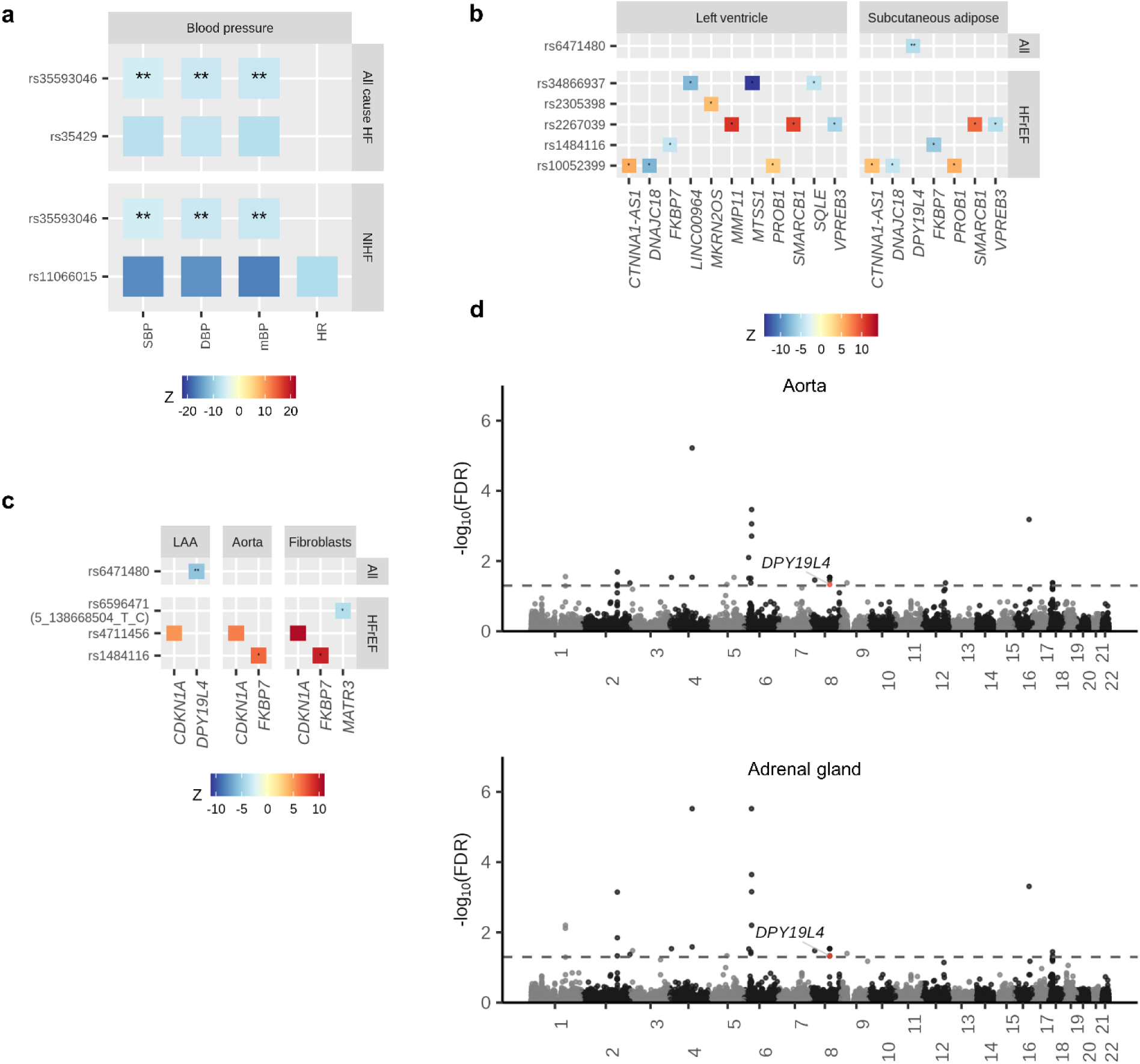
Downstream analysis of Japanese GWAS. **(a)** Lead variants identified in our Japanese GWAS of each HF phenotype are searched in previously reported Japanese GWAS of cardiovascular-related phenotypes. The color indicates the Z value of the corresponding Japanese GWAS. **(b and c)** Lead variants identified by each HF phenotype Japanese GWAS are searched in GTEx eQTL **(b)** and sQTL **(c)**. The color indicates the Z value of the corresponding eQTL or sQTL. rs6596471, which was strongly correlated with lead variant chr5:138668504:T:C (r^2^ > 0.8) is used as proxy variant since chr5:138668504:T:C was not found in GTEx database. **(d)** Manhattan plots of H-MAGMA using aorta or adrenal gland Hi-C datasets. -log_10_(FDR) on the *y*-axis are shown against the genomic positions (hg19) on the *x*-axis. Genes that have not been identified in previous TWAS, proteome-wide MR, or gene analysis are shown in red. ** indicates novel loci, and * indicates loci previously reported only in MTAG and not in GWAS. eQTL, expression quantitative trait loci; GWAS, genome-wide association study; HF, heart failure; HFpEF, heart failure with preserved ejection fraction; HFrEF, heart failure with reduced ejection fraction; H-MAGMA, Hi-C coupled MAGMA; LAA, left atrial appendage; MTAG, multi-trait analysis of GWAS; NIHF; non-ischemic heart failure; sQTL, splicing quantitative trait loci.

**Supplementary Fig. 5.**
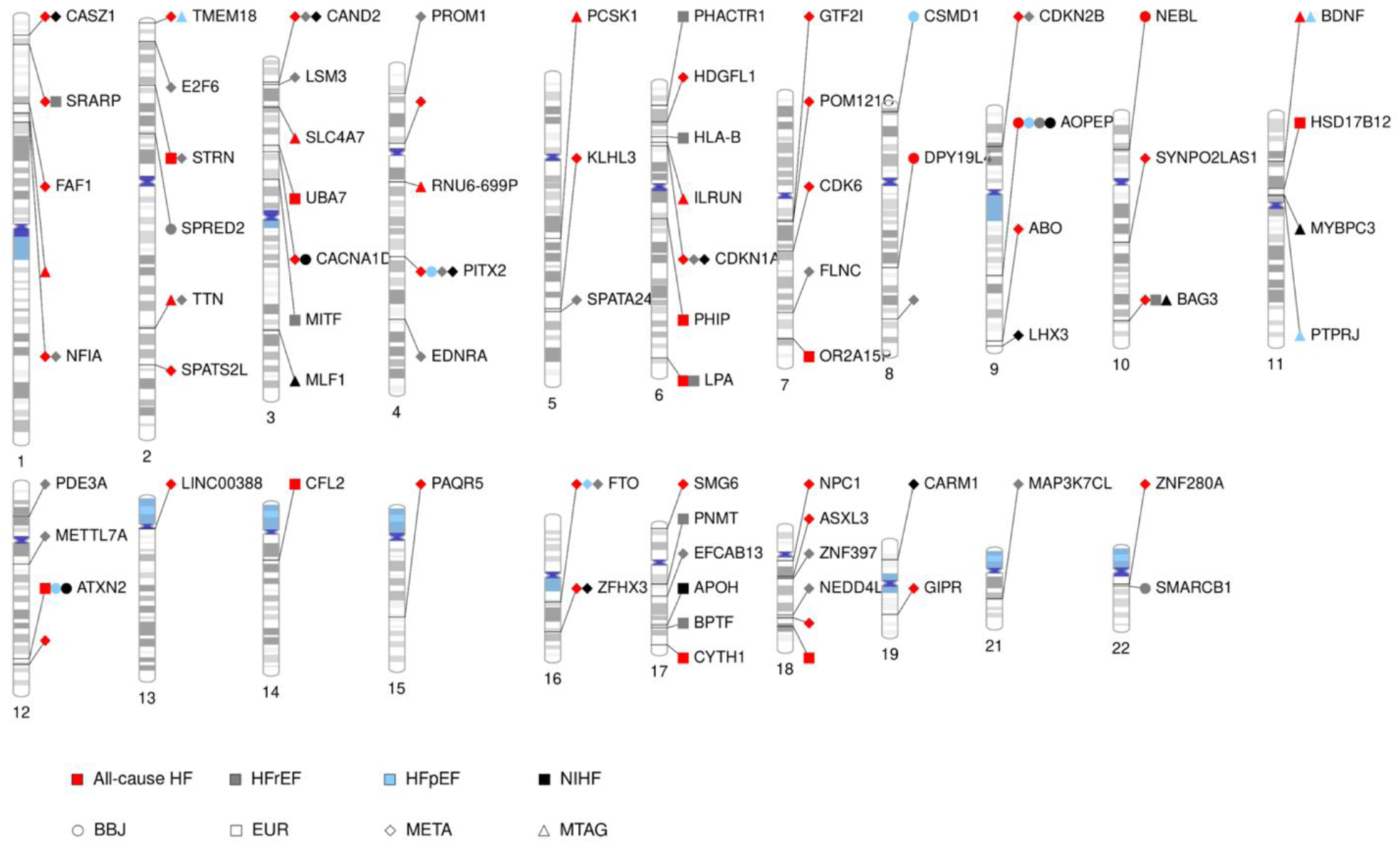
Phenogram of genome-wide significant loci for HF and its subtypes. Indicated gene names are the prioritized gene when one gene is prioritized by our methods, the nearest gene when multiple genes are prioritized equally, and the blank when none is prioritized and nearest gene is RNA gene. The shapes indicate HF subtypes; circles, all-cause HF; rectangles, HFrEF; triangles, HFpEF; and diamonds, non-ischemic HF. Colors indicate HF and its subtypes; red, BioBank Japan (BBJ); gray, European (EUR); light green, trans-ancestry meta-analysis (META); and black, multi-trait analysis of GWAS (MTAG). Cytobands and annotations are based on GRCh37/hg19. Gene names were annotated if prioritized by at least 3 methods.

**Supplementary Fig. 6.**
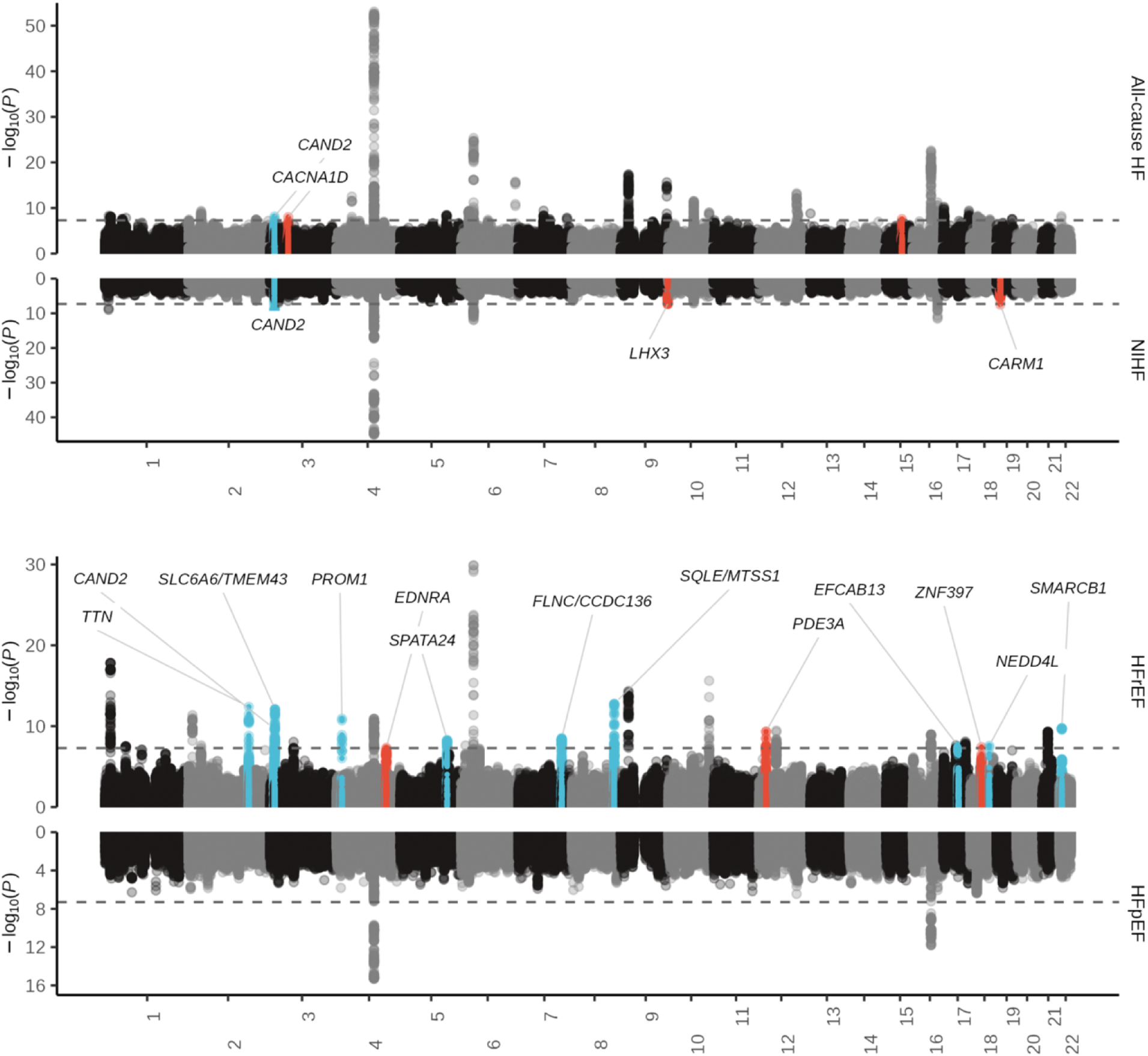
Results of the cross-ancestry meta-analysis. Miami plot of trans-ancestry meta-analysis. -log_10_(*P* value) on the *y*-axis is shown against the genomic positions (hg19) on the *x*-axis. Association signals that reached a genome-wide significance level (P < 5 × 10^-8^) are shown in red if new loci and in blue if previously reported only in MTAG but not in GWAS. GWAS, genome-wide association study; HF, heart failure; HFrEF, heart failure with reduced ejection fraction; HFpEF, heart failure with preserved ejection fraction; MTAG, multi-trait analysis of GWAS.

**Supplementary Fig. 7.**
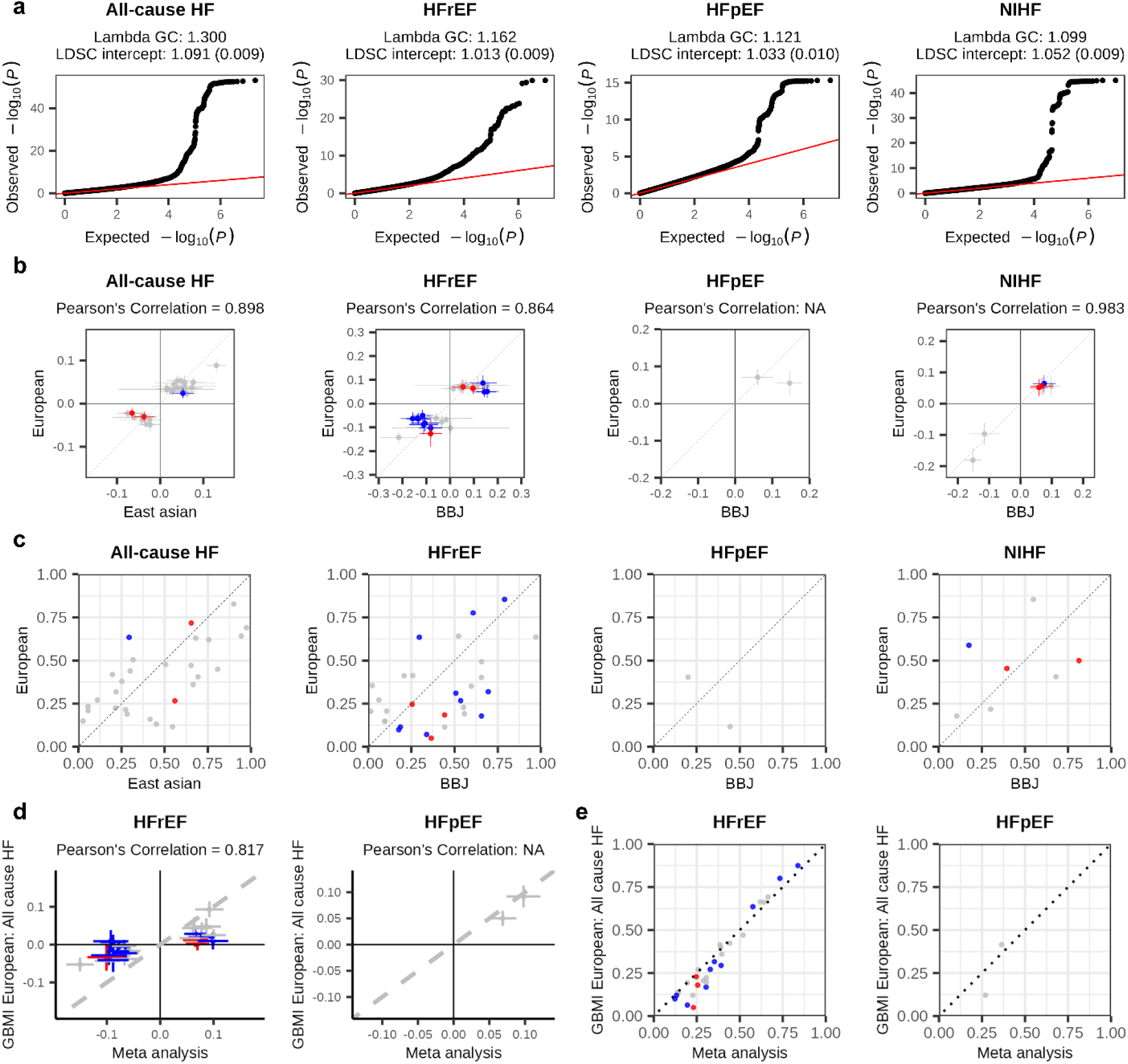
QQ plot and comparison of allele frequencies and allelic effects identified in the cross-ancestry meta-analysis. **(a)** QQ plot for each HF phenotype in the cross-ancestry meta-analysis. Two-sided *P* values were calculated using a logistic regression model. **(b** and **c)** Comparison of allelic effects **(b)** and allele frequency **(c)** between East Asian GWAS and European GWAS. Effect size with its 95%CI or allele frequency of East Asian (for all-cause HF) or BBJ (other HF phenotypes) GWAS are shown in the *x-*axis, and those of European are shown in the *y-*axis. **(d** and **e)** Comparison of allelic effects between the cross-ancestry meta-analysis (HFrEF or HFpEF) and non-overlapping European GWAS (GBMI European all-cause HF). Effect size with its 95%CI (**d**) or allele frequencies (**e**) of the cross-ancestry meta-analysis are shown in the *x*-axis, and those of European GWAS are shown in the *y-*axis. Points are shown in red if new loci and in blue if previously reported only in MTAG but not in GWAS. CI, confidence interval; GWAS, genome-wide association study; HF, heart failure; HFrEF, heart failure with reduced ejection fraction; HFpEF, heart failure with preserved ejection fraction; MTAG, multi-trait analysis of GWAS; QQ, quantile-quantile.

**Supplementary Fig. 8.**
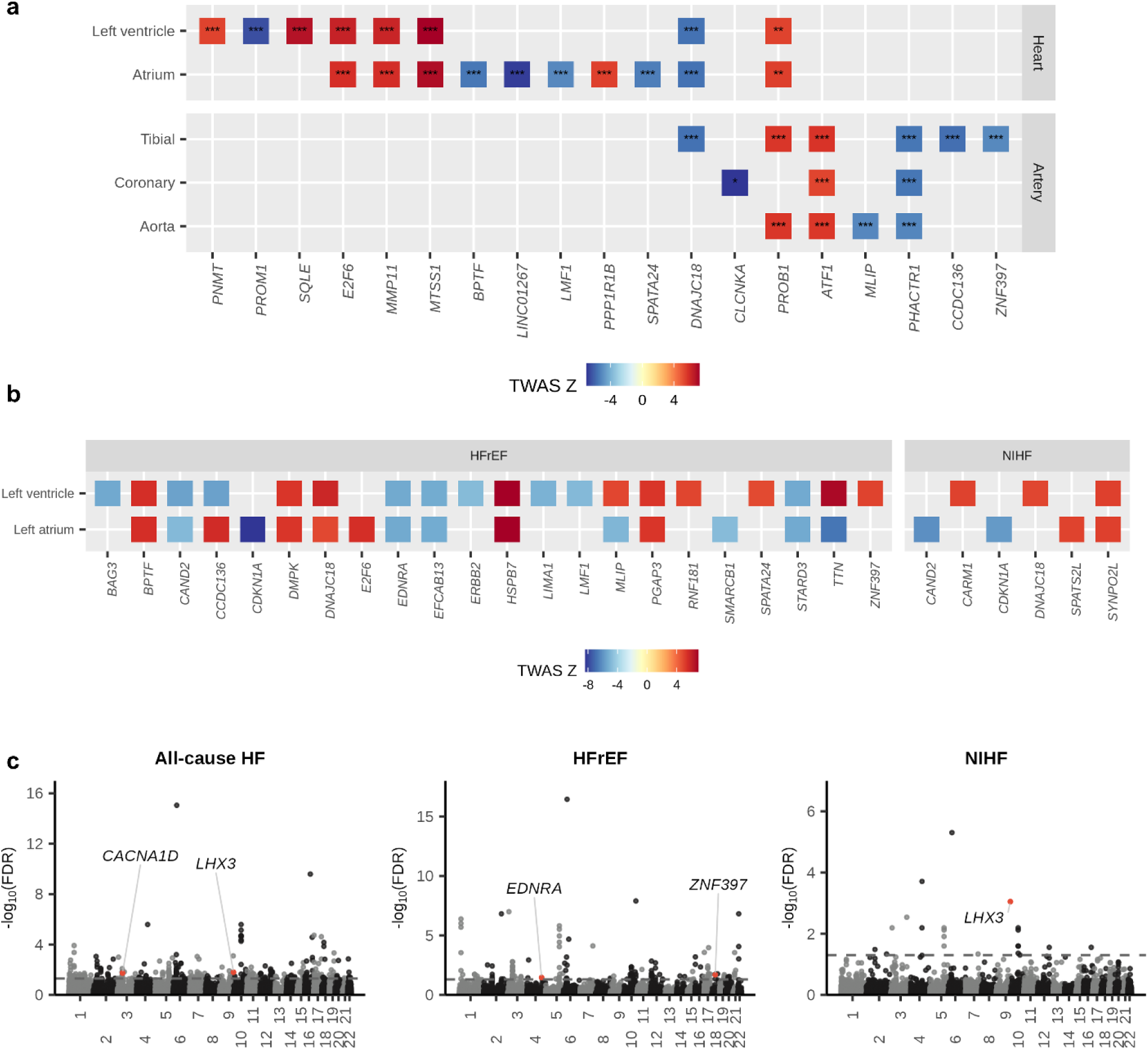
Downstream analysis of the cross-ancestry meta-analysis. **(a)** Heatmap of the TWAS of HF (all-cause HF and HF subtypes) showing transcriptome-wide significant associations with supporting evidence from colocalization. Colored squares TWAS significant associations based on two-sided *P* values after multiple testing corrections (P < 1.55 × 10^−6^). * indicate PP4 0.75-0.8, ** 0.8-0.9, *** 0.9-1.0. **(b)** Heatmap of the Splicing-TWAS HF showing splicing-wide significant associations. Colored squares splicing-TWAS significant associations based on two-sided *P* values after multiple testing corrections. Genes are presented on the *x*-axis, and tissue types are on the *y*-axis in both **(a)** and **(b)**. **(c)** Manhattan plots of MAGMA for all-cause HF, HFrEF, and non-ischemic heart failure. -log_10_(FDR) on the *y*-axis are shown against the genomic positions (hg19) on the *x*-axis. Genes that have not been identified in previous TWAS, proteome-wide MR, or gene analysis are shown in red. HF, heart failure; TWAS, transcriptome-wide association studies.

**Supplementary Fig. 9.**
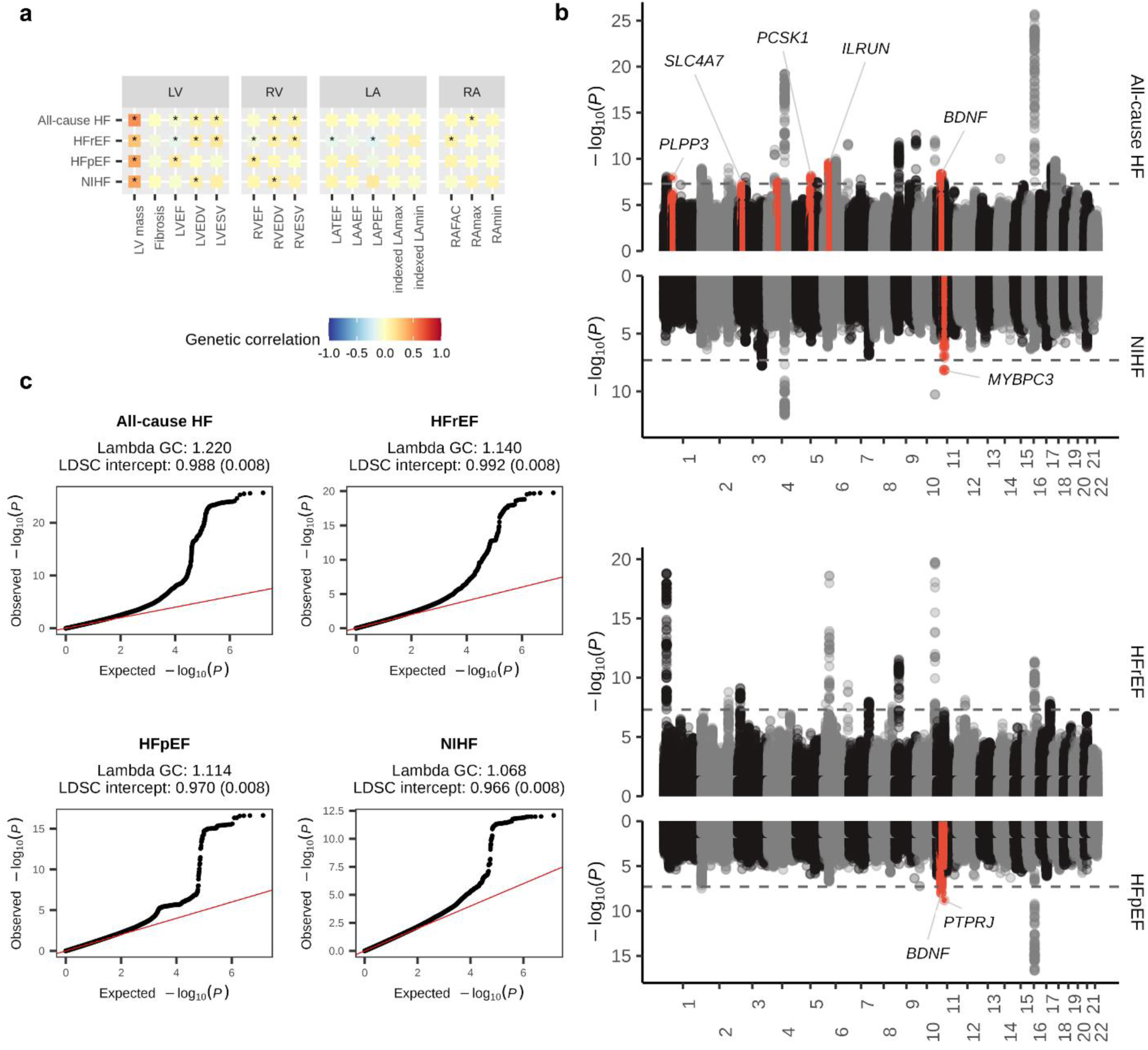
Results of multi-trait analysis of GWAS. **(a)** Genetic correlation between HF and cardiac MRI-derived parameters. Significant genetic correlations (P <0.05) are marked with an asterisk. Two-sided *P* values were calculated using linkage disequilibrium score regression. **(b)** Miami plot of MTAG. -log_10_(*P* value) on the *y*-axis are shown against the genomic positions (hg19) on the *x*-axis. Association signals that reached a genome-wide significance level (P < 5 × 10^-8^) are shown in red if new loci. **(c)** QQ plot for each HF phenotype in MTAG. GWAS, genome-wide association study; HF, heart failure; HFpEF, heart failure with preserved ejection fraction; HFrEF, heart failure with reduced ejection fraction; MTAG, multi-trait analysis of GWAS; NIHF, non-ischemic heart failure; QQ, quantile-quantile.

**Supplementary Fig. 10.**
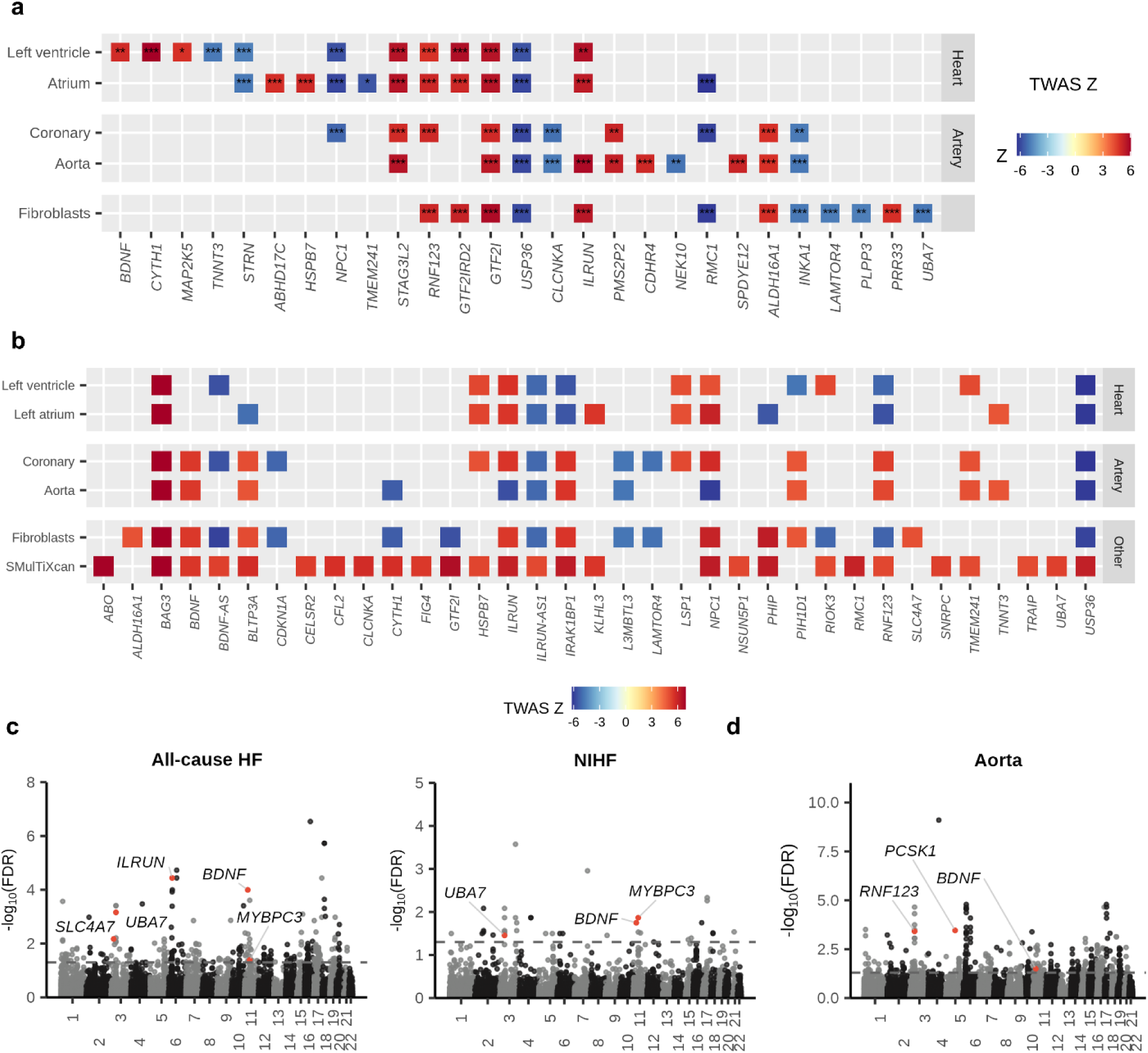
Downstream analysis of multi-trait analysis of GWAS. **(a)** Heatmap of the TWAS showing transcriptome-wide significant associations with supporting evidence from colocalization. Colored squares TWAS significant associations based on two-sided *P* values after multiple testing corrections (P < 1.55 × 10^−6^). * indicate PP4 0.75-0.8, ** 0.8-0.9, *** 0.9-1.0. **(b)** Heatmap of the Splicing-TWAS HF showing splicing-wide significant associations. Colored squares splicing-TWAS significant associations based on two-sided *P* values after multiple testing corrections. Genes are presented on the *x*-axis, and tissue types are on the *y*-axis in both **(a)** and **(b)**. **(c and d)** Manhattan plots of MAGMA for all-cause HF or non-ischemic heart failure **(c)**, and H-MAGMA using aorta Hi-C datasets in all-cause HF**(d)**. - log_10_(FDR) on the *y*-axis are shown against the genomic positions (hg19) on the *x*-axis. Genes that have not been identified in previous TWAS, proteome-wide MR, or gene analysis are shown in red. HF, heart failure; TWAS, transcriptome-wide association studies.

**Supplementary Fig. 11.**
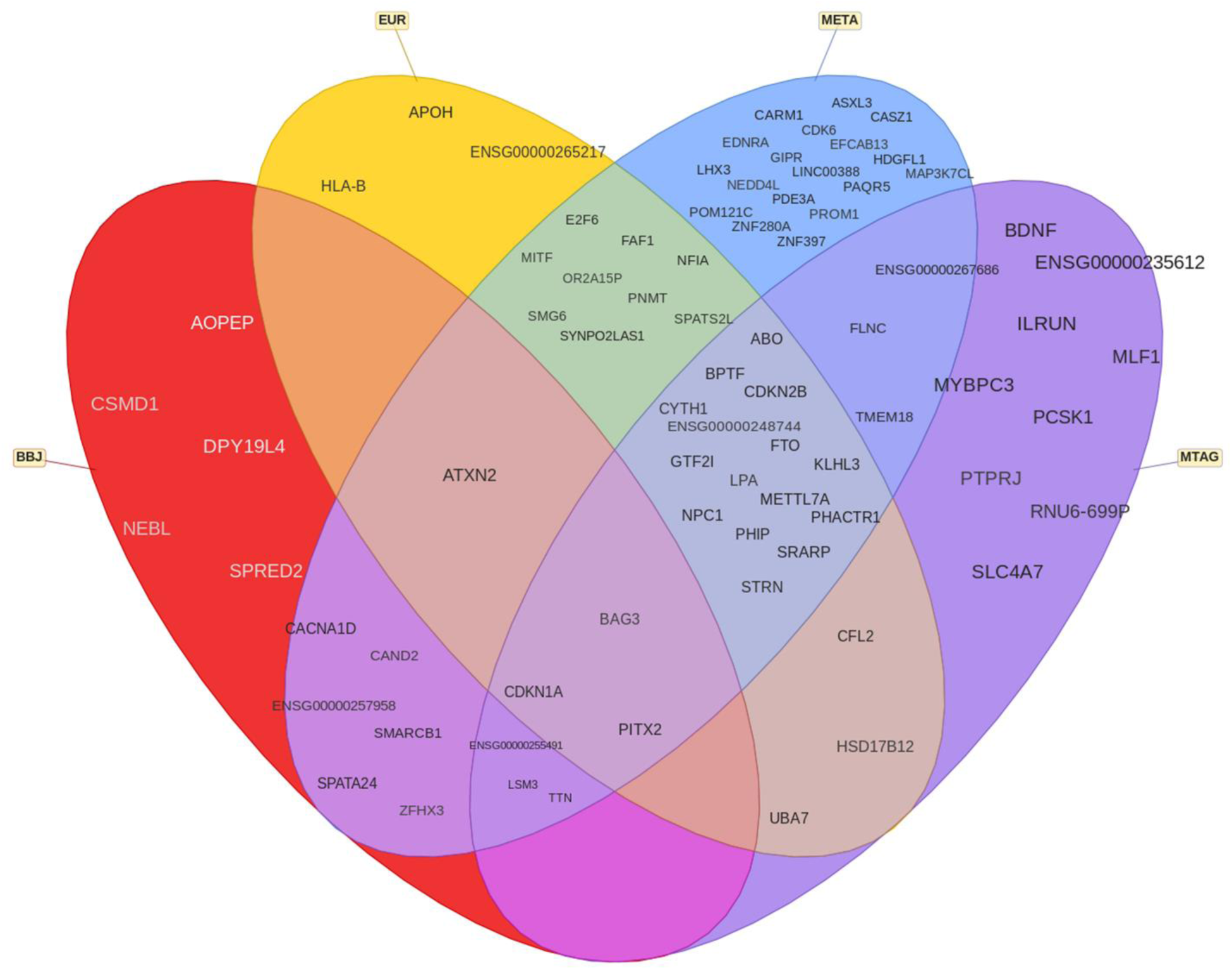
Benn diagram for genes identified by each analysis. BBJ, BioBank Japan; EUR, European; META, meta-analysis; MTAG, multi-trait analysis of GWAS.

**Supplementary Fig. 12.**
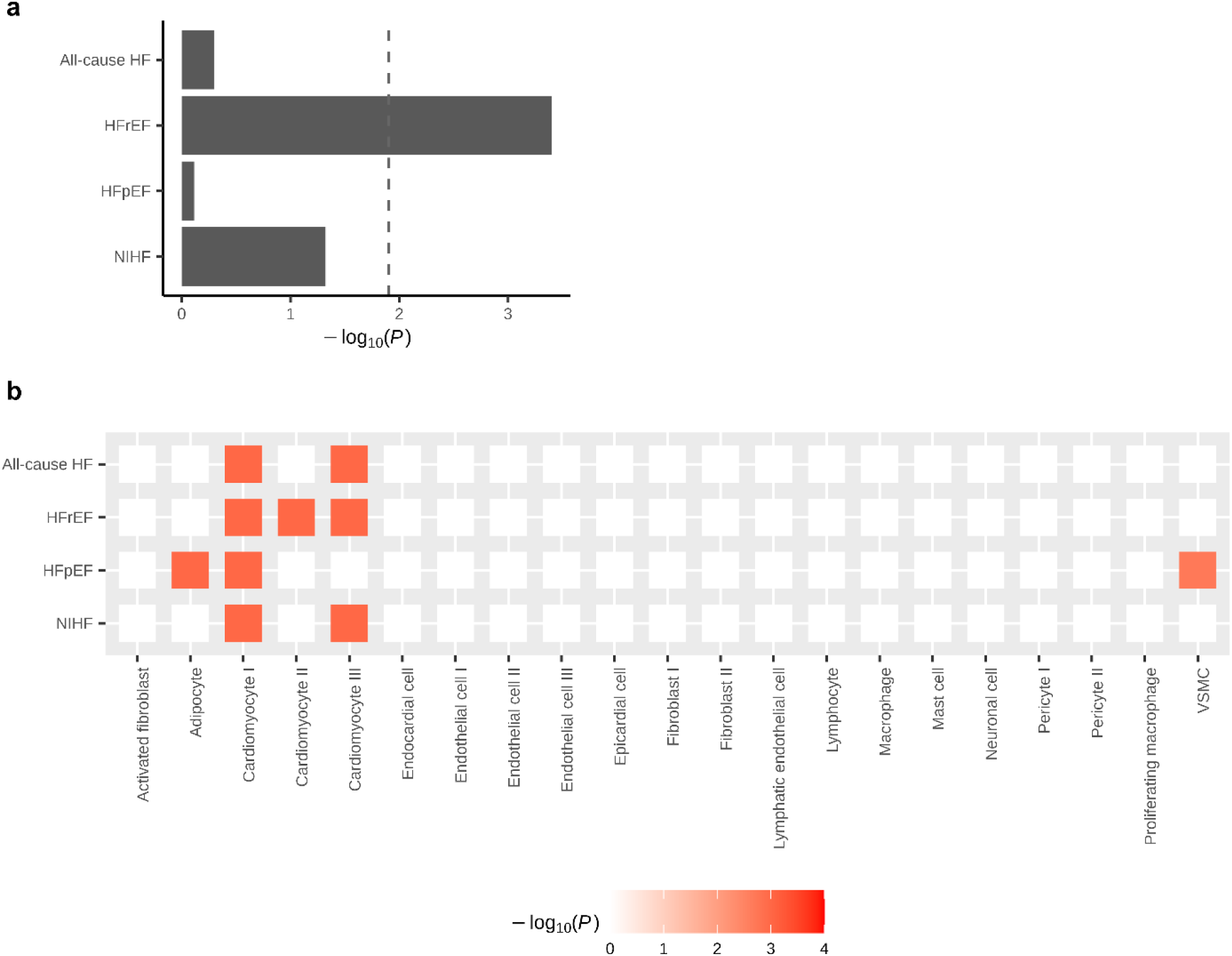
miRNA enrichment analysis and single-cell enrichment analysis. **(a)** MIGWAS results of the four HF phenotypes. The overall contribution of miRNA-target gene network to the traits through the tissue-naïve approach. An enrichment signal is shown by - log_10_(*P*_MIGWAS_). **(b)** Heatmaps depicting each cell type-disease association for HF phenotypes. Heatmap colors denote uncorrected *P* value of cell-type-disease association evaluated using scDRS. Only significant associations are shown.

**Supplementary Fig. 13.**
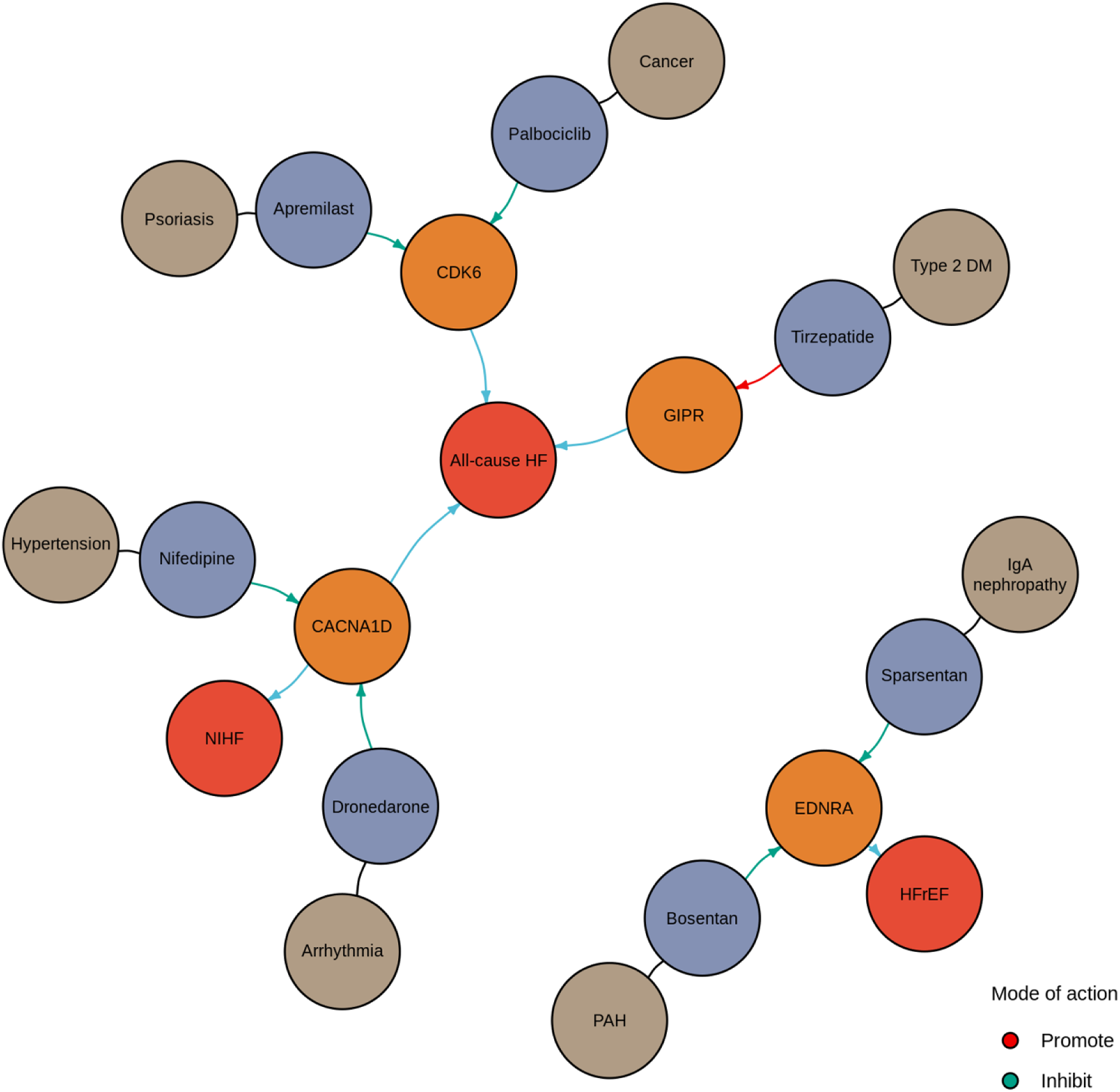
Candidate drugs linked to disease susceptibility loci for HF. Dark blue indicates HF subtypes; orange gene; purple medications; and brown diseases.

**Supplementary Fig. 14.**
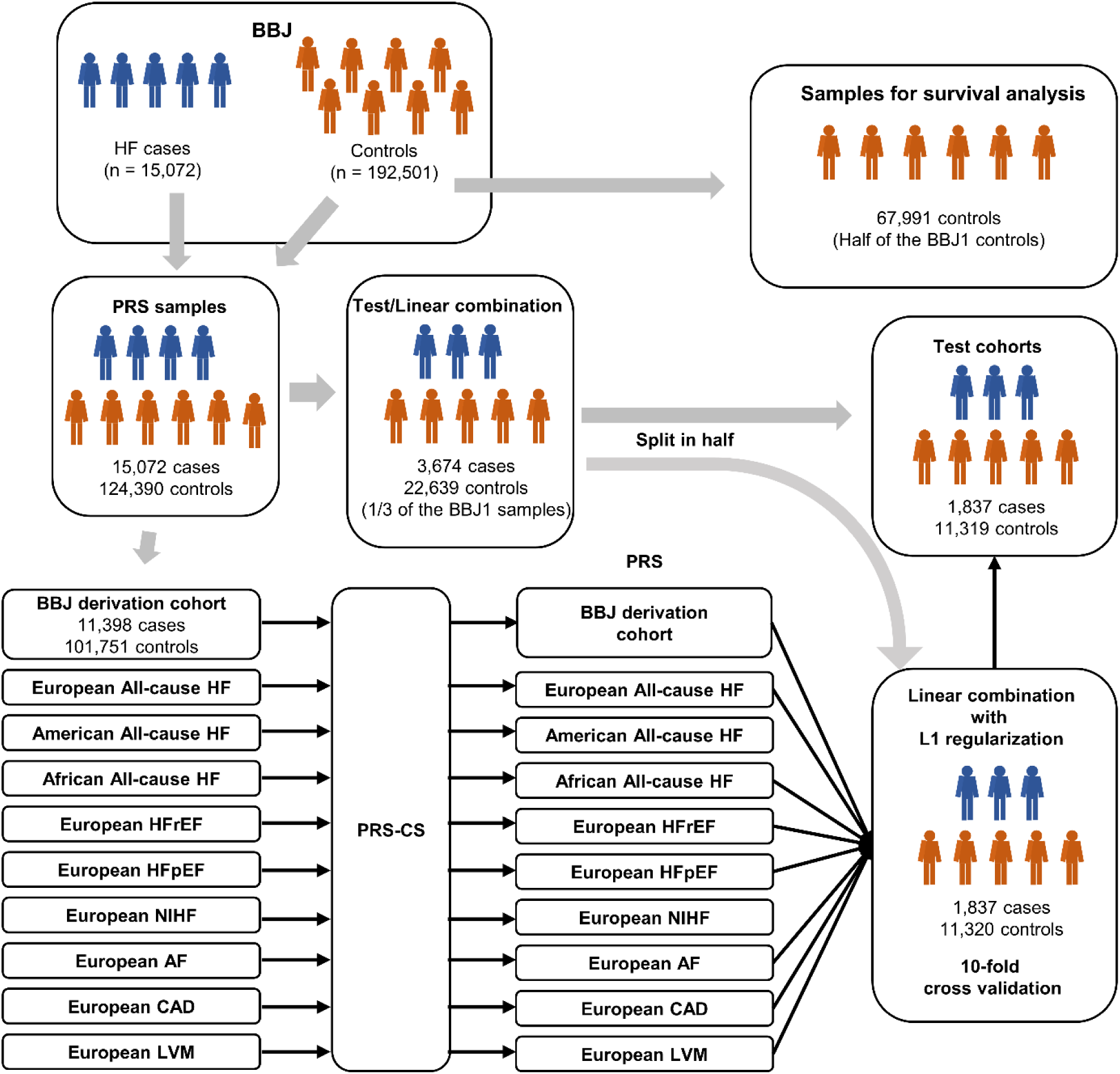
Analytical scheme for PRS development. Schematic representation for derivation, cross-validation, performance testing of HF-PRS in the independent test cohort, and survival analysis for HF-PRS.

**Supplementary Fig. 15.**
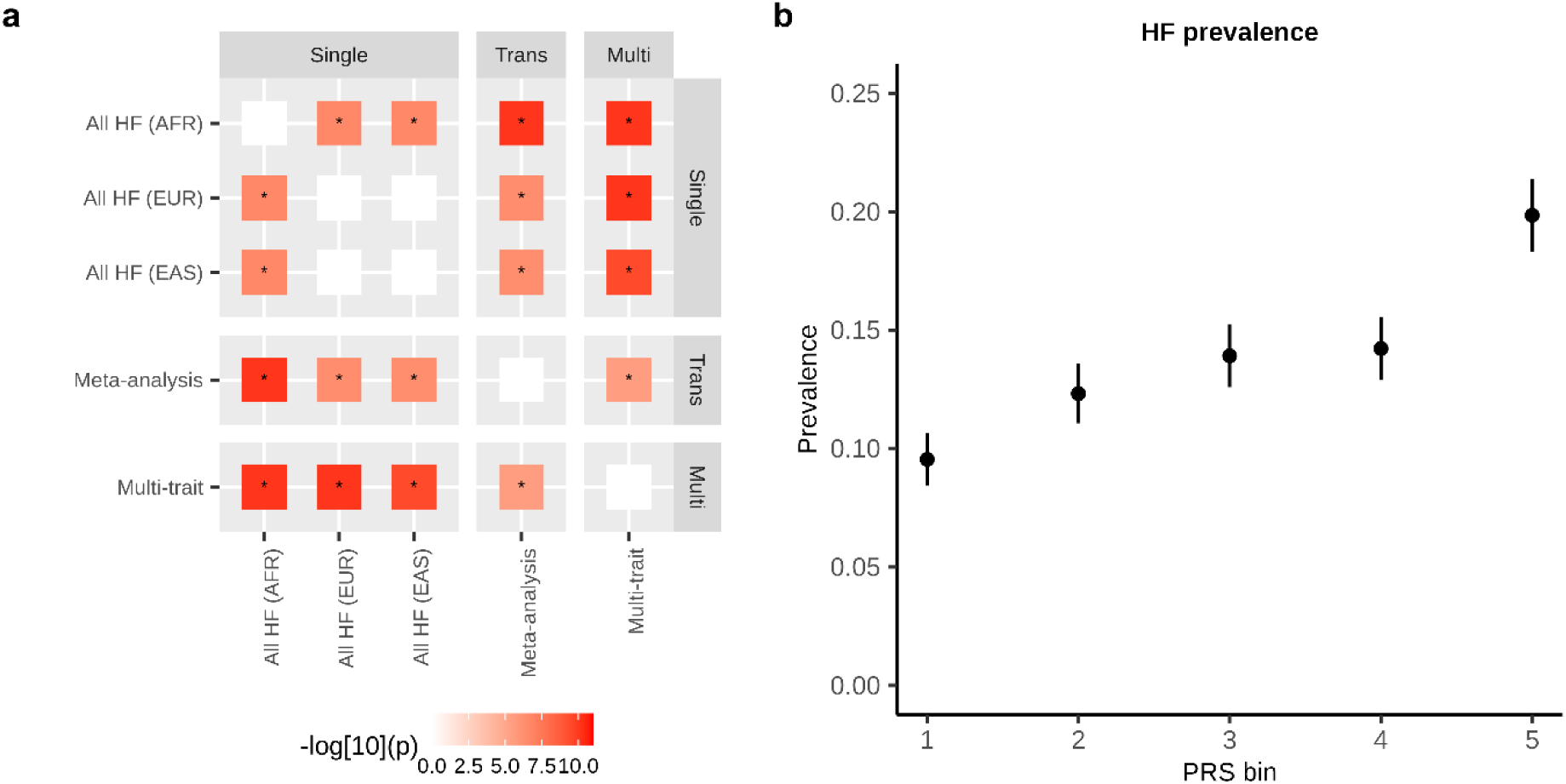
PRS performance. **(a)** Pairwise comparison of PRS performance. Distribution of the pairwise difference of Nagelkerke’s pseudo-R^2^ (Δ pseudo-R^2^: Pseudo R^2^_Score_ Y – Pseudo R^2^_Score_ X, X, and Y are found at the axis) between each pair of PRS models. The distributions were obtained by bootstrapping 5.0 × 10^4^ times. Two-sided bootstrap *P* values were calculated by counting the number of Δ pseudo-R^2^ ≤ 0 or Δ pseudo-R^2^ > 0 and then multiplying the lower value by the minimum estimated *P* value (2 * 1 / (5.0 × 10^4^) = 4 × 10^−5^: two-sided). The significance was set at P = 5.0 × 10−3 (0.05/10). **(b)** PRS distribution and HF prevalence. Prevalence of HF based on the HF-PRS deciles in each combination of GWAS. The number of individuals in each decile is 2,631-2,632. Data are presented as medians and 95% CI. PRS, polygenic risk score; CI, confidence interval.

